# Multimodal Machine Learning for Predicting Outcomes in the PASS-01 Trial of Systemic Therapy for Metastatic Pancreatic Cancer

**DOI:** 10.64898/2026.08.24.26360900

**Authors:** Wei Quan, David Henault, Amy Zhang, Gun Ho Jang, Syeda Mariam Hasnain, Daniela Bevacqua, Yangqing Deng, Eugenia Flores-Figueroa, Kewei Ni, Nicholas Light, Julie M. Wilson, Anna Dodd, Erica S. Tsang, Daniel A. King, Amber N. Habowski, Kenneth Yu, Kimberly Perez, Andrew J. Aguirre, Eileen M. O’Reilly, Brian M. Wolpin, Trevor J. Pugh, David A. Tuveson, Elizabeth M. Jaffee, Steven Gallinger, Grainne O’Kane, Faiyaz Notta, Jennifer J. Knox, Robert C. Grant

**Affiliations:** Princess Margaret Cancer Centre, University Health Network, Toronto, Ontario, Canada; Ontario Institute for Cancer Research, Toronto, Ontario, Canada; Department of Biostatistics, University Health Network, Toronto, Ontario, Canada; Northwell, New Hyde Park, New York, United States; Lustgarten Foundation Pancreatic Cancer Research Laboratory, Cold Spring Harbor Laboratory, Cold Spring Harbor, New York, United States; Memorial Sloan Kettering Cancer Center and Weill Cornell Medical College, New York, United States; Dana Farber Cancer Institute and Harvard Medical School, Boston, Massachusetts, United States; Sidney Kimmel Comprehensive Cancer Center Johns Hopkins University School of Medicine, Baltimore, Maryland, United States; St. Vincent’s University Hospital and School of Medicine, University College Dublin, Dublin, Ireland

## Abstract

**Purpose:** Modified FOLFIRINOX (FFX) and gemcitabine plus nab-paclitaxel (GNP) are standard first-line treatments for metastatic pancreatic ductal adenocarcinoma (PDAC), but no validated biomarker guides treatment selection. We developed MULTIPL, a multimodal machine learning system, and established the PASS-01 Challenge to benchmark prognostic and predictive biomarkers.

**Patients and Methods:** MULTIPL was trained in the COMPASS study (N=268), integrating clinical, digitized histopathology, whole-genome, and RNA-seq data. MULTIPL, PurIST, hENT1 expression, and HRDetect were evaluated in the PASS-01 trial, a randomized phase II trial of FFX versus GNP (N=160), within the Challenge. The primary endpoint was differential treatment benefit measured by concordance-for-benefit for progression-free survival.

**Results:** MULTIPL had the highest concordance index for OS among individually evaluated biomarkers (0.595; 95% confidence interval [CI], 0.55–0.65) and separated high-versus low-risk patients (hazard ratio, 1.62; 95% CI, 1.13–2.33; *P*=0.009). Patients recommended for GNP by MULTIPL had significantly longer OS with GNP than with FFX (hazard ratio, 0.47; 95% CI, 0.28–0.82; *P*=0.007), whereas patients recommended for FFX had similar OS between treatments. Interpretability analysis of MULTIPL in COMPASS identified *KDM6A* alterations and *SSTR1* expression as prognostic biomarkers, which were validated in PASS-01. However, none of the tested biomarkers significantly predicted differential treatment benefit in the PASS-01 Challenge.

**Conclusion:** MULTIPL demonstrated robust prognostic performance in external validation, identified a subgroup enriched for benefit from GNP, and enabled discovery and validation of prognostic biomarkers in metastatic PDAC. However, no biomarker met the primary endpoint for differential treatment benefit, underscoring the value of the PASS-01 Challenge.

**Translational Relevance:** Several biomarkers have been proposed to guide first-line treatment selection in metastatic pancreatic cancer, but none are validated from randomized data. We developed MULTIPL, a multimodal machine-learning model that integrates clinical, histopathologic, genomic, and transcriptomic data from the observational COMPASS study. In parallel, we launched the PASS-01 Challenge to evaluate biomarkers in a randomized trial of modified FOLFIRINOX versus gemcitabine plus nab-paclitaxel to evaluate predictive and prognostic biomarkers. Neither MULTIPL nor the published biomarkers PurIST, hENT1, and HRDetect met the prespecified endpoint for predicting differential treatment benefit measured using concordance for benefit. MULTIPL nevertheless demonstrated prognostic capabilities and identified a subgroup with longer survival on gemcitabine plus nab-paclitaxel. Model interpretation also identified *KDM6A* alterations and *SSTR1* expression as prognostic biomarkers, which were validated in PASS-01. These findings demonstrate the potential of multimodal machine learning in pancreatic cancer and establish the PASS-01 Challenge as a randomized evaluation of biomarkers for treatment selection.

## Introduction

Pancreatic ductal adenocarcinoma (PDAC) is projected to become the second leading cause of cancer death by 2030(1). For patients with newly diagnosed metastatic disease, choosing among first-line regimens remains a central clinical problem. Modified FOLFIRINOX (fluorouracil, leucovorin, irinotecan, and oxaliplatin; FFX), NALIRIFOX (liposomal irinotecan, fluorouracil, leucovorin, and oxaliplatin), and gemcitabine plus nab-paclitaxel (GNP) are widely used first-line options(2,3). Across contemporary trials, these regimens yield a median overall survival (OS) of approximately one year(4–7). Yet population-level averages do not identify the regimen most likely to benefit an individual patient, because variation in clinical features and tumor biology may produce clinically meaningful differences in treatment effect.

Several biomarkers have been proposed to guide first-line treatment selection, including classical and basal-like transcriptomic subtypes, hENT1 expression, and homologous recombination deficiency(8–12). Most were developed and evaluated in observational cohorts, in which treatment selection is influenced by age, performance status, disease burden, and physician preference. Associations between biomarkers and treatment-specific outcomes may therefore reflect systematic differences in why patients received each treatment (confounding by indication), rather than true treatment-effect modification. Randomized trials provide the most rigorous setting for distinguishing prognostic biomarkers, which are associated with outcomes irrespective of treatment, from predictive biomarkers, which identify differential benefit between treatments.

The PASS-01 trial provides an opportunity to evaluate biomarkers within a randomized trial comparing FFX and GNP in metastatic PDAC(5). Pretreatment tumor biopsies were collected for digitized whole-slide histopathology images (WSIs) and, after laser-capture microdissection, for whole-genome sequencing (WGS) and transcriptome sequencing (WTS). The COMPASS study provides a complementary development cohort with similar multimodal profiling in an observational cohort of patients with advanced PDAC treated primarily with FFX or GNP(13). Together, these cohorts enable model development on observational data and independent evaluation on randomized trial data.

Using COMPASS, we developed MULTIPL, a multimodal machine-learning system that generates treatment-specific predictions of response and survival for FFX or GNP. In parallel, we established the PASS-01 Challenge, in which multimodal or individual biomarkers can be submitted for centralized performance evaluation using pre-specified metrics. We then benchmarked MULTIPL, PurIST, hENT1, and HRDetect in the PASS-01 Challenge and externally validated model-derived prognostic biomarkers.

## Methods

### Study oversight and reporting standards

The COMPASS and PASS-01 studies were approved by the relevant research ethics boards. All participants provided informed consent for clinical data collection, molecular profiling, and translational research in accordance with the governing trial protocols. MULTIPL was developed and evaluated according to TRIPOD+AI reporting principles(14).

### Cohorts

COMPASS (NCT02750657) was a prospective precision-medicine study of patients with advanced PDAC who were receiving first-line systemic therapy, selected by the treating physician and patient. The development cohort included patients treated with modified FFX or GNP who had prospective clinical annotation and available molecular or histopathological profiling. COMPASS was used for model development because it links pretreatment tumor biopsies to clinical outcomes, WGS, WTS, and WSI in a cohort with advanced PDAC(13,15).

PASS-01 (NCT04469556) was a multi-institutional randomized phase II trial of previously untreated metastatic PDAC. Patients were assigned to FFX or GNP and underwent pretreatment biospecimen collection with multimodal profiling, including WGS, WTS, and WSI(5).

### Features and targets

Clinical, treatment, and outcome data were collected prospectively within the COMPASS and PASS-01 protocols. Candidate clinical variables were selected before modeling based on availability, clinical relevance, and alignment with COMM-PACT consensus domains(16).

Pretreatment tumor biopsies underwent formalin fixation and were embedded in paraffin, stained with hematoxylin and eosin, and digitized at 40x magnification. Samples underwent laser-capture microdissection prior to nucleic acid extraction to enrich for the malignant epithelial compartment(17,18). Molecular features included somatic genomic alterations, mutational signatures, and normalized gene expression values processed using previously reported bioinformatics pipelines(5,13). Clinical, WGS, and WTS features were entered into models as tabular features. WSIs were converted into fixed-length embeddings using GigaPath(19,20).

Objective response (ORR) was defined as complete or partial response versus stable disease, progressive disease, or non-evaluable disease according to trial radiologic assessment. Overall survival (OS) was available in COMPASS and PASS-01. Progression-free survival (PFS) was available in PASS-01 but not in COMPASS.

### MULTIPL development

MULTIPL was developed in COMPASS to generate treatment-specific predictions for patients receiving FFX or GNP from the multimodal data including clinical, genomic, transcriptomic, and histopathological inputs. Separate models were trained within each treatment group for two binary endpoints: ORR and one-year OS. Patients censored before one year without an observed death were excluded. Because PFS was unavailable in COMPASS, an exploratory PFS score was derived from ORR and one-year OS predictions using a weighting function that anchored on OS.

Model development used repeated nested cross-validation in COMPASS. Ten random seeds were used, each generating a five-fold outer split. Within each training fold, preprocessing, imputation, feature selection, scaling, and hyperparameter tuning were performed without access to the held-out fold. Candidate learners included ElasticNet logistic regression(21), XGBoost(22), and TabPFN(23,24). Candidate integration strategies included early fusion of preprocessed features and late fusion of unimodal predictions. The optimal configuration was selected in cross-validation, and the final model was retrained on the full COMPASS cohort. For differential treatment effects analyses, patient-level treatment-effect scores were computed as the predicted outcome under FFX minus the predicted outcome under GNP.

### The PASS-01 Challenge

The PASS-01 Challenge (https://hpb-research.ca) is an open benchmarking initiative designed to evaluate prognostic and treatment-selection algorithms in randomized PDAC trial data.

Participants will receive baseline clinical, pathological, and molecular PASS-01 data without outcome labels. Participants will then submit patient-level predictions to the Challenge team for central evaluation in accordance with prespecified protocols. Participants will receive results within two weeks of submission, which can be published immediately.

The primary Challenge objective is to predict differential treatment effects of GNP and FFX on PFS in the per-protocol cohort, evaluated using the concordance for benefit (C4B) statistic(25). Submissions also include prognostic and treatment-effect predictions for ORR and OS, which are evaluated in the intention-to-treat population.

### Benchmark biomarkers

MULTIPL was compared with three prespecified, literature-derived biomarkers: PurIST subtype(11), hENT1 expression(12), and HRDetect(26). These biomarkers were selected because they represent distinct PDAC treatment-selection hypotheses and could be implemented using available PASS-01 molecular data. PurIST classified tumors as classical or basal-like based on RNA sequencing; classical cancers have been proposed to benefit more from FFX(9). hENT1 was assessed using *SLC29A1* RNA expression; high expression has been proposed to be associated with treatment response(12). HRDetect was used as a genomic scar measure of homologous recombination deficiency and as a platinum-based chemotherapy biomarker, with a cutoff of 0.7(26,27).

### Model interpretability analysis

Model interpretability analyses were performed to identify biological features contributing to MULTIPL predictions. SHapley Additive exPlanations (SHAP) values were computed for treatment-arm-specific one-year OS models in COMPASS to rank features by their contribution to model predictions(28). For DNA and RNA models, features were ranked by their mean absolute SHAP value. Top-ranked features were selected using the elbow approach, followed by association testing in COMPASS. Features meeting the false-discovery threshold of 0.1 were carried forward for PASS-01 validation using COMPASS-derived thresholds.

For histopathology, SHAP-selected GigaPath(20) embedding dimensions were not directly interpretable as biological variables. To assign biological context, top-ranked embedding features were associated with previously reported expert-annotated histopathological features(29), using Spearman correlations. Annotations included stromal phenotype, growth pattern, and WHO grade.

### Statistical analysis

Prediction of differential treatment effects was evaluated using the C4B metric(25) for ORR, adapted to survival outcomes for PFS and OS(25,30). A value of 0.5 is consistent with no treatment selection ability, while 1.0 indicates perfect ability to select the best treatment for each patient. Secondary exploratory analysis examined differences between treatment arms in biomarker-defined subpopulations using Cox proportional hazards regression. Predictions of ORR were evaluated using the area under the receiver operating characteristic curve (AUC), while predictions of OS and PFS were evaluated using Harrell’s concordance index(31). To evaluate the ability of predictions to separate high- and low-risk subpopulations, scores were dichotomized using COMPASS-derived thresholds at the median unless otherwise specified. Uncertainty was summarized using 95% confidence intervals estimated by bootstrap resampling. To promote broad applicability of the evaluated biomarkers, missing values for features were imputed using the medians for continuous features and modes for categorical features. This means, for example, where RNAseq was unavailable, patients were classified as classical for PurIST. Complete-case analysis was considered in secondary analyses. Statistical tests were two-sided.

## Results

### Cohorts and study design

In the COMPASS development cohort, clinical data and WGS were available for 268 patients, WTS for 253, and digitized WSI for 199. Complete multimodal profiles were available for 188 patients (Fig. 1A). ORR occurred in 41/144 (28.5%) of evaluable FFX-treated patients and 30/99 (30.3%) of evaluable GNP-treated patients. One-year OS occurred in 64/146 (43.8%) of FFX-treated patients and 29/99 (29.3%) of GNP-treated patients (Fig. 1B). Baseline characteristics differed between treatment groups, with older ages in the GNP arm (Table S1).

**Fig. 1:**
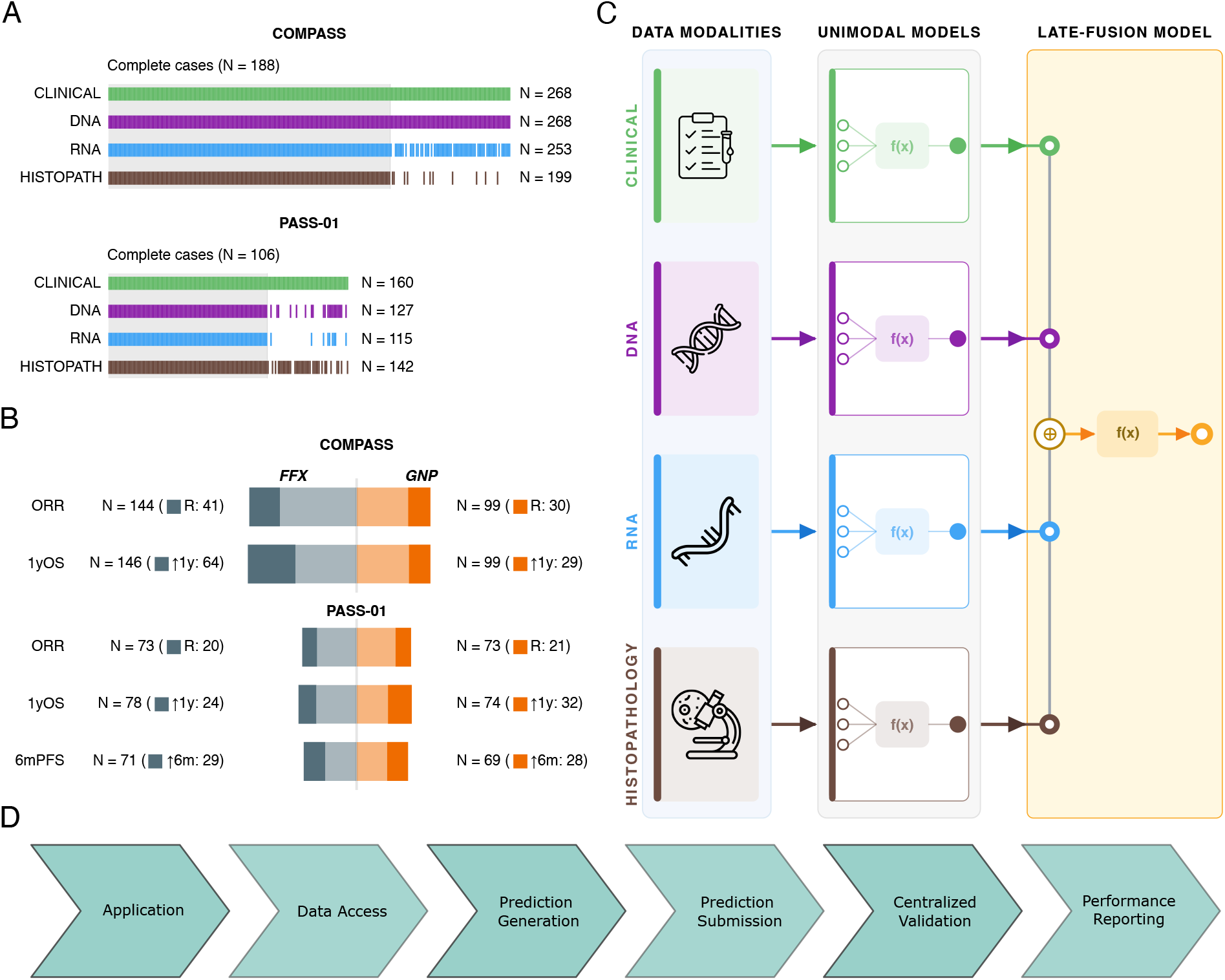
Cohorts, MULTIPL architecture, and PASS-01 Challenge workflow. **A**, Availability of clinical, genomic, transcriptomic, and histopathologic data in COMPASS and PASS-01. Bars indicate the number of patients in each modality; numbers above each cohort indicate the number of patients with complete four-modality profiles. **B**, Clinical endpoints by treatment arm. Bars indicate evaluable patients, with the number meeting each endpoint annotated. Endpoints included objective response rate, one-year overall survival, and 6-month progression-free survival, which was available only in PASS-01. **C**, MULTIPL architecture. Modality-specific models generate predictions that are integrated by a late-fusion meta-learner. **D**, PASS-01 Challenge workflow from team application through centralized evaluation and performance reporting. Challenge details are available at https://hpb-research.ca/. FFX, modified FOLFIRINOX; GNP, gemcitabine plus nab-paclitaxel; ORR, objective response rate; OS, overall survival; PFS, progression-free survival.

The PASS-01 validation cohort included 160 randomized patients in the intention-to-treat population, with 80 assigned to FFX and 80 to GNP; the per-protocol population used for the primary differential-treatment-effect endpoint (C4B) comprised 140 patients (71 FFX, 69 GNP) (Table S1). Clinical data were available for all patients, WGS for 127, WTS for 115, and digitized WSI for 142. Complete four-modality profiles were available for 106 patients (Fig. 1A). ORR was evaluable in 73 patients per arm, with responses in 20 (27.4%) FFX-treated and 21 (28.8%) GNP-treated patients. For time-to-event outcomes, 63 deaths occurred in the FFX arm and 53 in the GNP arm, and 68 progression-or-death events occurred in each arm.

### Development of MULTIPL in COMPASS

We first developed treatment-specific models for one-year OS and objective response rate (ORR) in COMPASS. Across repeated nested cross-validation, performance varied by treatment, data modality, learner, and fusion strategy; no single modality or model family consistently dominated (Fig. 2; Fig. S1). Predictions from clinical, genomic, transcriptomic, and histopathologic models were only partly correlated, indicating that the modalities captured nonredundant information (Fig. S2).

**Fig. 2:**
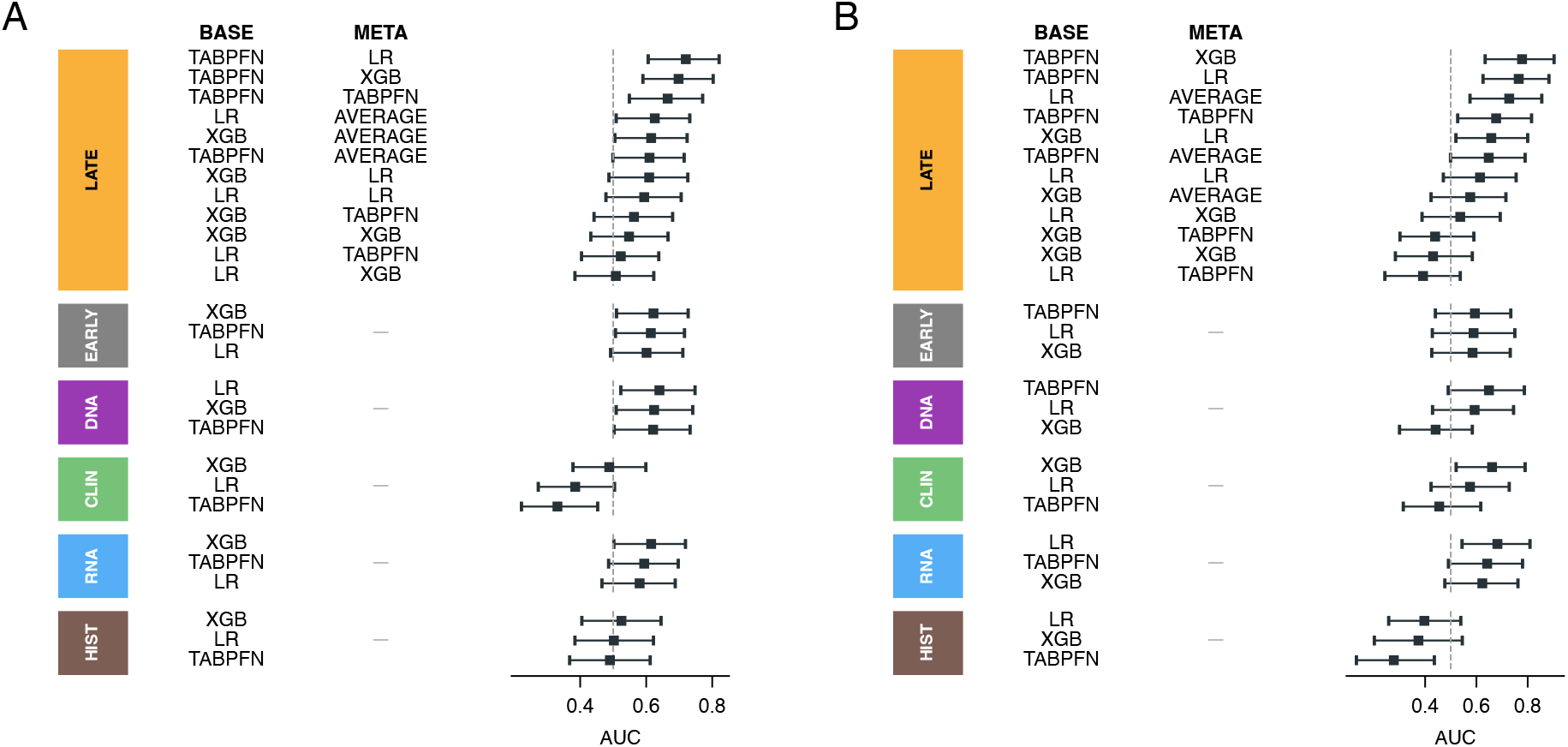
Development of MULTIPL for one-year overall survival prediction in COMPASS. Cross-validated performance of candidate models in patients treated with FFX (**A**) and GNP (**B**). Points indicate area under the receiver operating characteristic curve; horizontal lines indicate 95% bootstrap confidence intervals. Models are grouped by fusion strategy and unimodal data type. For late-fusion models, combinations of base-learner and meta-learner are shown. Analyses were restricted to patients with complete multimodal data. AUC, area under the receiver operating characteristic curve; CLIN, clinical; HIST, histopathology; FFX, modified FOLFIRINOX; GNP, gemcitabine plus nab-paclitaxel.

The best one-year OS model combined TabPFN unimodal predictions with a logistic regression late-fusion model. The cross-validated AUC was 0.730 (95% CI, 0.644–0.814) for FFX and 0.739 (95% CI, 0.621–0.848) for GNP. ORR models showed acceptable discrimination for FFX (AUC, 0.705; 95% CI, 0.594–0.801) but weaker discrimination for GNP (AUC, 0.633; 95% CI, 0.507–0.754). The final models were retrained in the full COMPASS development set and locked before PASS-01 outcomes were accessed.

In exploratory COMPASS analyses, the MULTIPL OS score identified an FFX-favoring subgroup with a numerical advantage for FFX over GNP (median OS, 11.6 versus 9.1 months; hazard ratio [HR] for GNP versus FFX, 1.45; 95% CI, 0.98–2.13; P=0.061) but did not identify a corresponding GNP advantage in the GNP-favoring subgroup (HR, 1.31; 95% CI, 0.89–1.93; P=0.172; Fig. S3). Because treatment allocation was not randomized in COMPASS, these analyses were considered descriptive.

### Model interpretation in COMPASS

We examined the biological features underlying MULTIPL predictions. SHAP attribution was applied to the treatment-specific one-year OS models in COMPASS, and features were ranked by their mean absolute contribution to model output (Fig. 3A).

**Fig. 3:**
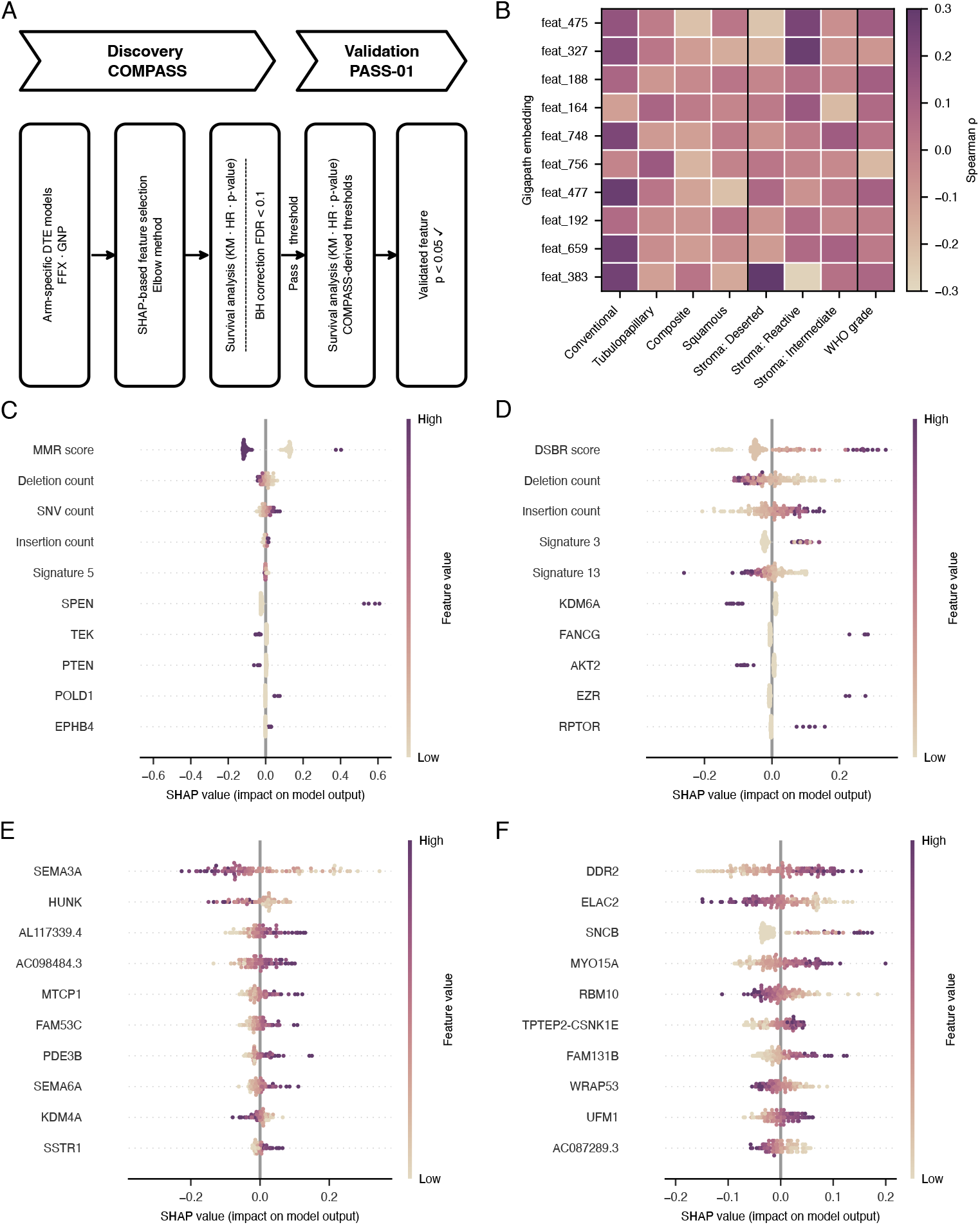
Interpretability analysis of MULTIPL. **A**, Analysis workflow. SHAP attribution was applied to treatment-specific one-year overall survival models, followed by feature selection in COMPASS and validation in COMPASS and PASS-01. **B**, Spearman correlations between SHAP-selected GigaPath histopathology embedding dimensions and expert-annotated histopathologic features. **C–F**, SHAP summary plots for the DNA model in the GNP arm (**C**), the DNA model in the FFX arm (**D**), the RNA model in the GNP arm (**E**), and the RNA model in the FFX arm (**F**). Features are ranked by mean absolute SHAP value; each point represents one patient, and color indicates feature value. FFX, modified FOLFIRINOX; GNP, gemcitabine plus nab-paclitaxel; SHAP, Shapley additive explanations.

Histopathology embeddings were interpreted by correlating SHAP-selected GigaPath features with expert-annotated morphological characteristics. The strongest associations were with stromal phenotype and growth pattern (Fig. 3B). Selected embeddings distinguished deserted from reactive stroma and conventional from nonconventional growth, suggesting that the histopathology model captured tumor architecture and stromal organization beyond conventional grading.

The DNA models recovered features related to DNA repair. In the FFX model, Signature 3, which is linked to homologous recombination(32), was associated with longer OS in COMPASS, whereas Signature 18, linked to damage by reactive oxygen species, and *KDM6A* alterations were associated with shorter OS (Fig. 3D; Table S2). In the GNP DNA model, only a higher mismatch-repair score(26) was associated with shorter OS in COMPASS (Table S4). RNA models identified additional distinct treatment-associated features. Ten and eight transcripts were associated with the FFX and GNP models, respectively (Fig. 3E-F; Table S3, S5). DNA and RNA features associated with OS in COMPASS were carried forward for validation in PASS-01 when they met the prespecified false-discovery threshold of 0.1.

### Evaluation of MULTIPL in PASS-01

We next evaluated locked MULTIPL predictions in PASS-01 in accordance with the pre-specified Challenge process. The primary endpoint, prediction of differential PFS benefit, measured by C4B, was 0.572 (95% CI, 0.38–0.67; P=0.329). C4B was also nonsignificant for OS (0.517; 95% CI, 0.46–0.71; P=0.794) and objective response (0.491; 95% CI, 0.36–0.62; P=0.886). Thus, MULTIPL did not meet the prespecified endpoint for individualized treatment-selection performance.

Exploratory treatment-stratified analyses suggested MULTIPL identified a subset of patients who benefit from GNP more than FFX. Among patients where MULTIPL recommended GNP, median OS was 13.0 months with GNP and 8.7 months with FFX (HR for GNP versus FFX, 0.47; 95% CI, 0.28–0.82; P=0.007; Fig. 4A). Among patients where MULTIPL recommended FFX, median OS was similar between treatment groups (8.9 months with GNP versus 9.0 months with FFX; HR, 0.88; 95% CI, 0.52–1.49; P=0.638). The treatment-by-MULTIPL interaction was P=0.205. The corresponding PFS differences were smaller (Fig. 4C) and nonsignificant for ORR (Fig. 4B).

**Fig. 4:**
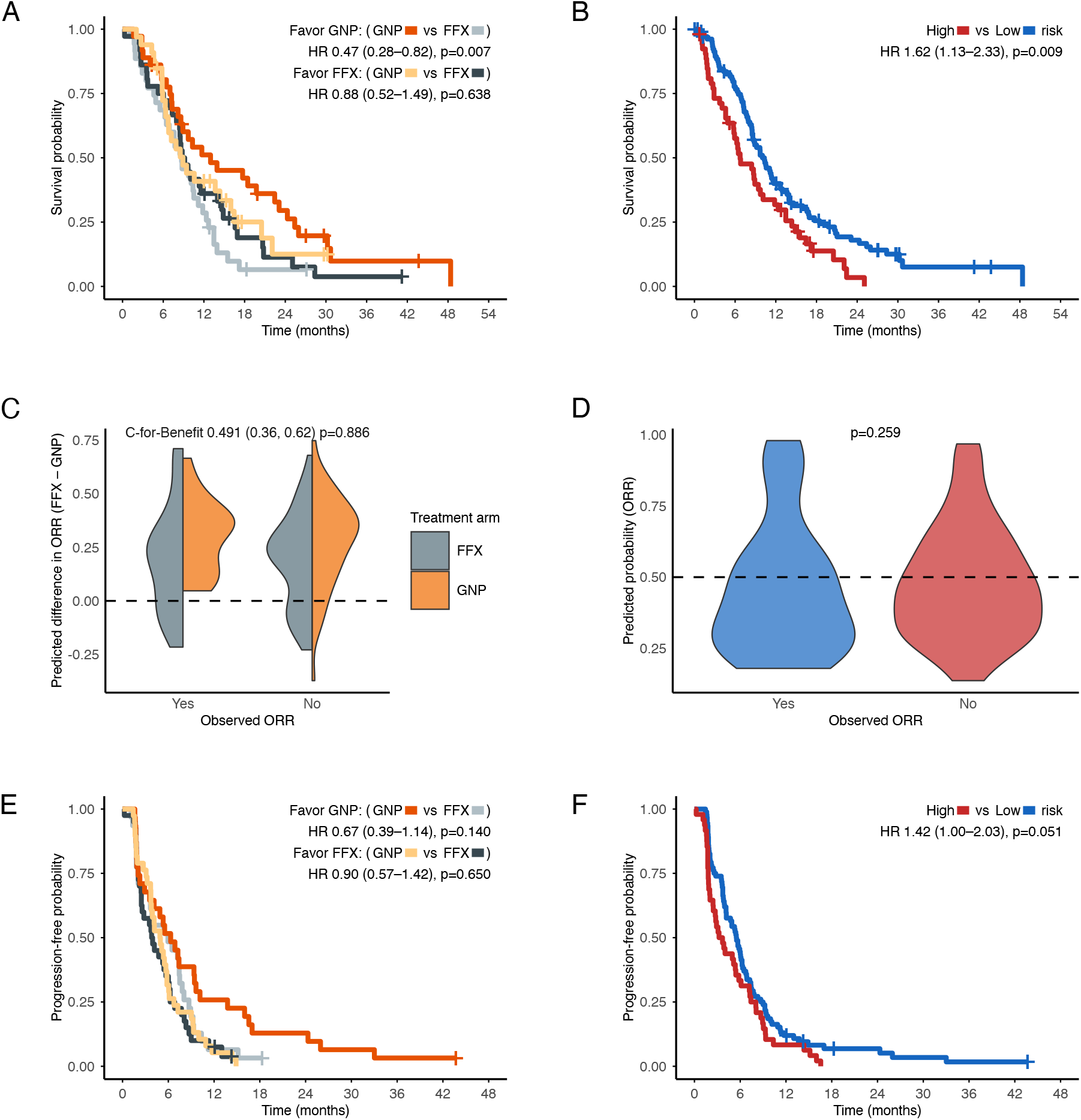
MULTIPL performance in the PASS-01 Challenge. **A–C**, Differential treatment effects analyses, for overall survival (**A**), objective response (**B**), and progression-free survival (**C**). Treatment-effect scores were calculated as the predicted outcome with FFX minus the predicted outcome with GNP; positive values favor FFX. Kaplan–Meier curves are stratified by randomized treatment within model-defined treatment-recommendation groups. **D–F**, Prognostic analyses for overall survival (**D**), objective response (**E**), and progression-free survival (**F**). Risk groups were defined using COMPASS-derived thresholds. Hazard ratios compare GNP with FFX in treatment-effect analyses and high-risk with low-risk in prognostic analyses. FFX, modified FOLFIRINOX; GNP, gemcitabine plus nab-paclitaxel; ORR, objective response; OS, overall survival; PFS, progression-free survival.

MULTIPL demonstrated prognostic discrimination for OS, with a c-index of 0.595 (95% CI, 0.55–0.65; P<0.001). Patients classified as high risk had shorter OS than those classified as low risk, with median OS of 6.7 versus 10.0 months (HR, 1.62; 95% CI, 1.13–2.33; P=0.009; Fig. 4D). The exploratory PFS predictions also retained prognostic discrimination. The PFS c-index was 0.563 (95% CI, 0.51–0.62; P=0.021), and median PFS was 3.4 months in the high-risk group compared with 5.5 months in the low-risk group (HR, 1.42; 95% CI, 1.00–2.03; P=0.051; Fig. 4F). By contrast, MULTIPL did not predict ORR (AUC, 0.440; 95% CI, 0.34–0.55; P=0.273; Fig. 4E).

### Validation of benchmark submissions to PASS-01

We then evaluated three prespecified literature-derived biomarkers: PurIST, hENT1 expression, and HRDetect. None met the primary endpoint for differential PFS benefit. PFS C4B was 0.534 for PurIST (95% CI, 0.41–0.66; P=0.621), 0.447 for hENT1 (95% CI, 0.29–0.57; P=0.461), and 0.499 for HRDetect (95% CI, 0.29–0.59; P=0.987; Fig. 5A). C4B estimates for OS and ORR were also nonsignificant.

**Fig. 5:**
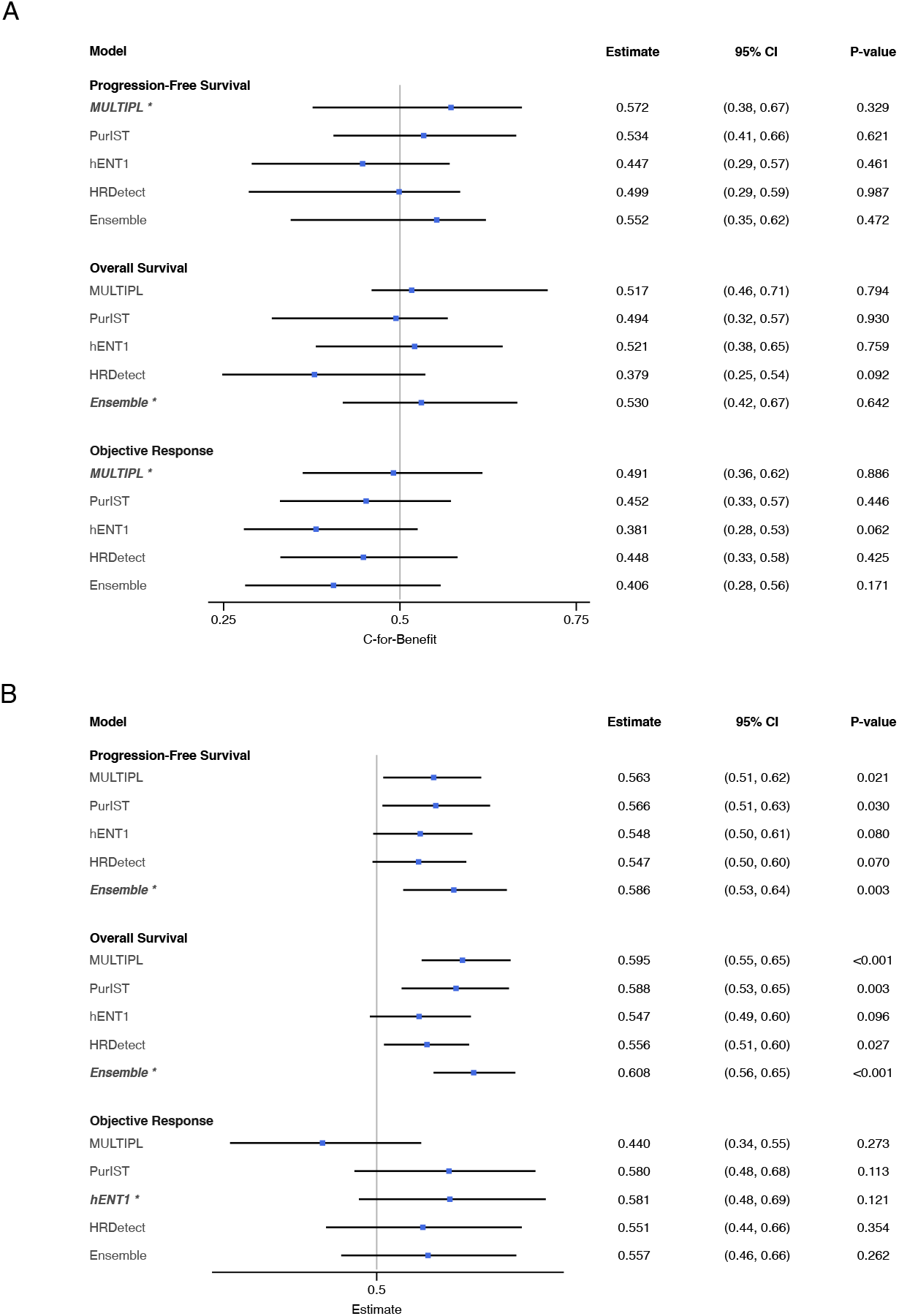
PASS-01 Challenge performance of MULTIPL and benchmark biomarkers. **A**, Differential treatment effects performance for progression-free survival, overall survival, and objective response, measured by concordance for benefit. **B**, Prognostic performance for the same endpoints, measured by Harrell concordance index for survival outcomes and area under the receiver operating characteristic curve for objective response. Points indicate performance estimates; horizontal lines indicate 95% bootstrap confidence intervals. A value of 0.5 indicates no discrimination. Models include MULTIPL, PurIST, hENT1, HRDetect, and a rank-average ensemble. Asterisks denote the highest estimate for each endpoint. AUC, area under the receiver operating characteristic curve; CI, confidence interval.

Exploratory analyses within treatment arms identified additional associations. Among patients with classical tumors defined by PurIST, outcomes were superior with GNP versus FFX for OS (median OS 13.0 versus 9.4 mo; HR, 0.63; 95% CI, 0.40–0.97; P=0.037), in contrast to previous observational cohorts; however, this difference was not observed for PFS. High hENT1 expression identified a small GNP-favoring subgroup with longer OS on GNP, but this subgroup contained only 13 patients (Fig. S5). HRDetect-defined treatment groups were highly imbalanced, with few HRDetect-high patients, since PASS-01 excluded patients with known or suspected germline *BRCA1*, *BRCA2*, and *PALB2* mutations (Fig. S6).

The benchmark biomarkers retained varying degrees of prognostic information. PurIST showed the strongest individual benchmark performance, with c-indices of 0.588 for OS (95% CI, 0.53– 0.65; P=0.003) and 0.566 for PFS (95% CI, 0.51–0.63; P=0.030; Fig. 5B). HRDetect showed weaker OS discrimination (c-index, 0.556; 95% CI, 0.51–0.60; P=0.027), whereas hENT1 did not significantly discriminate either survival endpoint. None of the benchmark biomarkers significantly predicted ORR.

### Integrative analysis

We next examined whether the top-ranked prognostic biomarkers, MULTIPL and PurIST, captured overlapping prognostic information. Their one-year OS predictions were essentially uncorrelated in PASS-01 (r=0.029; P=0.755; Fig. 6A). Patients classified as high risk by both models had the shortest survival, with a median OS of 4.9 months, compared with 12.7 months among patients classified as low risk by both models (HR, 2.92; 95% CI, 1.62–5.28; P<0.001; Fig. 6B). Survival differed across all four concordant and discordant risk groups (log-rank P=0.003). Among patients with cancers classified as basal-like by PurIST, MULTIPL was prognostic (HR, 2.00; 95% CI, 1.01–3.94; P=0.042), with a similar numeric trend among patients with PurIST classical cancers (HR, 1.52; 95% CI 0.89–2.58; P=0.121). These results indicate that MULTIPL learned prognostic signals that were partly independent of established transcriptomic subtyping. Furthermore, this supports integrating multiple biomarkers into an ensemble in the PASS-01 Challenge.

**Fig. 6:**
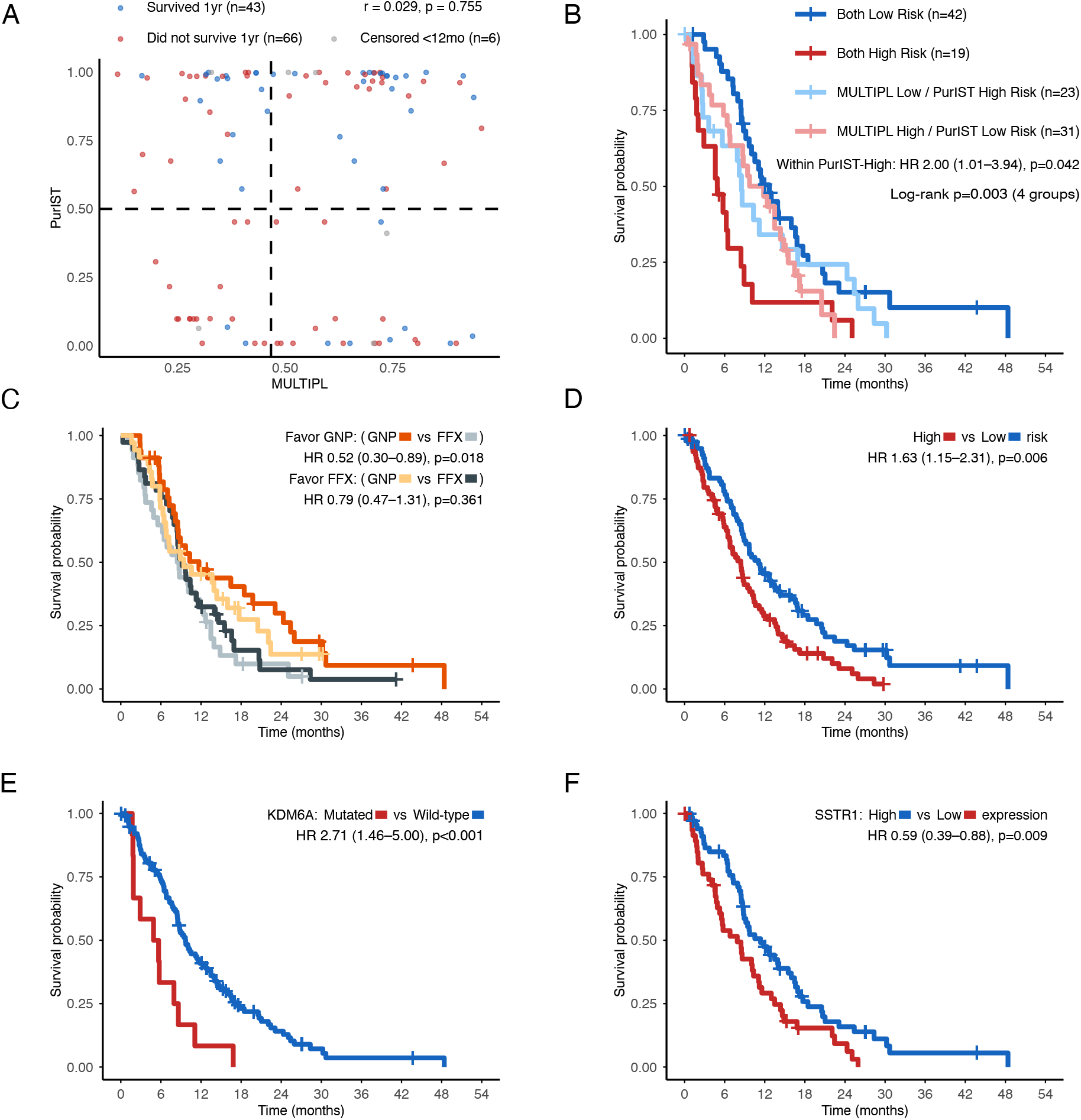
Integrative analyses and validation of model-derived biomarkers in PASS-01. **A**, Association between MULTIPL predictions of one-year overall survival and PurIST classification. Points are colored by observed one-year survival; dashed lines indicate the MULTIPL and PurIST classification thresholds. **B**, Overall survival according to concordant and discordant MULTIPL and PurIST risk classifications. **C**, Overall survival by randomized treatment within treatment-recommendation groups defined by the rank-average ensemble. **D**, Overall survival by ensemble prognostic risk group. **E**, **F**, Overall survival according to KDM6A mutation status (E) and SSTR1 expression (F). Hazard ratios compare mutated with wild-type tumors in A, high with low expression in B, GNP with FFX in E, and high with low risk in F. FFX, modified FOLFIRINOX; GNP, gemcitabine plus nab-paclitaxel; OS, overall survival.

We evaluated a rank-average ensemble of MULTIPL, PurIST, hENT1, and HRDetect. The ensemble did not significantly predict differential treatment benefit for PFS, OS, or ORR (Fig. 5A). Similar to the pattern observed with MULTIPL and PurIST, among patients classified as favoring GNP, median OS was 11.7 months with GNP and 8.6 months with FFX (HR, 0.52; 95% CI, 0.30–0.89; P=0.018; Fig. 6C). However, the ensemble achieved the highest overall prognostic performance, with c-indices of 0.608 for OS (95% CI, 0.56–0.65; P<0.001) and 0.586 for PFS (95% CI, 0.53–0.64; P=0.003; Fig. 5B). Ensemble-defined high-risk patients had shorter OS than low-risk patients (median, 8.4 versus 11.1 months; HR, 1.63; 95% CI, 1.15–2.31; P=0.006; Fig. 6D) and shorter PFS (median, 3.7 versus 6.1 months; HR, 1.62; 95% CI, 1.15– 2.28; P=0.006; Fig. S7).

### Validation of MULTIPL-derived biomarkers

Finally, we tested whether individual genomic biomarkers nominated through MULTIPL interpretation validated in PASS-01. Two features retained the direction and magnitude of association observed in COMPASS. Patients with *KDM6A*-altered tumors had shorter OS than those with wild-type tumors. Median OS was 5.3 months in the altered group and 9.7 months in the wild-type group (HR, 2.71; 95% CI, 1.46–5.00; P<0.001; Fig. 6E). High *SSTR1* expression was associated with longer OS. Median OS was 11.4 months in the high-expression group and 7.9 months in the low-expression group (HR, 0.59; 95% CI, 0.39–0.88; P=0.009; Fig. 6F). Other DNA and RNA features nominated in COMPASS did not reproduce individually in PASS-01 (Tables S2-S5).

## Discussion

Optimally selecting first-line chemotherapy remains one of the most important unanswered clinical questions in metastatic PDAC. Although FFX and GNP produce relatively similar outcomes at the population level, clinicians currently lack validated biomarkers to guide treatment selection for individual patients. In this study, we developed a multimodal machine-learning system using clinical, histopathological, genomic, and transcriptomic data, and evaluated it in PASS-01, the first randomized trial designed to compare biomarker-guided treatment selection between FFX and GNP. While MULTIPL demonstrated validated prognostic performance and identified biologically plausible prognostic biomarkers, neither MULTIPL nor three previously proposed biomarkers significantly predicted differential treatment benefit according to the prespecified primary endpoint in the PASS-01 Challenge.

The PASS-01 Challenge establishes a framework for prospective evaluation of treatment-selection algorithms using randomized clinical trial data, locked predictions, prespecified evaluation metrics, and centralized validation. This addresses a longstanding limitation in biomarker development, in which competing models are typically evaluated across different retrospective observational cohorts using varied endpoints and analytical methods. By providing a common benchmark, PASS-01 enables direct comparison of multimodal models and molecular biomarkers, and establishes a mechanism for rigorously evaluating future algorithms. Promising biomarkers that could not be evaluated in this study have been proposed for treatment selection(33), and we encourage submission to the Challenge.

Perhaps our most important finding is the negative primary result. Numerous biomarkers have been proposed to guide chemotherapy selection in PDAC, including molecular subtypes, hENT1 expression, and homologous recombination deficiency, yet all have been developed and evaluated in observational datasets, which cannot separate predictive effects from confounding by indication. The PASS-01 Challenge removes this source of bias through randomized treatment allocation. Under these conditions, and in light of the eligibility criteria including exclusion of suspected or known *BRCA1/2* and *PALB2* pathogenic variants, none of the evaluated biomarkers demonstrated statistically significant treatment-selection performance. These results demonstrate the difficulty in identifying predictive biomarkers and the importance of randomization in their evaluation.

MULTIPL demonstrated prognostic validity in PASS-01. After development in observational COMPASS data, the locked model achieved the highest OS discrimination among the individual biomarkers evaluated and separated patients into clinically distinct risk groups. MULTIPL and PurIST were essentially uncorrelated, and patients classified as high risk by both were associated with the shortest survival. These findings indicate that biomarkers capture complementary prognostic signals and support their integrative approaches for risk stratification.

Although the primary treatment-selection endpoint was negative, exploratory analyses identified a subgroup predicted by MULTIPL to benefit from GNP that demonstrated substantially longer overall survival when randomized to GNP. This result illustrates a distinction between subgroup analysis and differential treatment effects, which test different hypotheses. Subgroup analysis tested whether GNP improves outcomes over FFX within a biomarker-defined population, whereas concordance for benefit tested whether MULTIPL correctly ranks differential treatment effects across all patients. A negative concordance-for-benefit result therefore does not exclude a subgroup effect, but the observed GNP-favoring signal was exploratory and requires additional confirmation.

PASS-01 did not confirm the observational hypothesis that classical PDAC preferentially benefits from FFX(9,10,13). In the PASS-01 randomized data, classical tumors instead had significantly longer OS with GNP, although this pattern was not observed for PFS and PurIST did not significantly predict differential treatment benefit. Earlier reports favoring FFX in classical disease were based on nonrandomized cohorts, in which treatment choice reflected patient fitness and disease severity. Preferential assignment of patients with more aggressive basal-like disease to GNP could therefore bias outcomes against GNP, creating an apparent FFX-specific benefit in classical tumors without true treatment-effect modification. Randomization in PASS-01 removes this confounding by indication and supports interpreting classical–basal subtypes as prognostic, rather than a validated biomarker for selecting FFX over GNP.

While we did not observe a differential treatment benefit for treatment selection using HRDetect, the PASS-01 trial excluded patients with known or strongly suspected germline pathogenic variants in *BRCA1*, *BRCA2*, and *PALB2*, so this result should be interpreted cautiously. Few cancers were HRDetect positive, and prior evidence strongly supports platinum-based chemotherapy in pancreatic cancers with homologous recombination deficiency(26,34).

Our findings support the potential of multimodal learning while defining its limits. Clinical, histopathological, genomic, and transcriptomic models generated only modestly correlated predictions, indicating that each modality may capture partly distinct signals. In COMPASS, late-fusion MULTIPL achieved the numerically highest one-year OS performance in both treatment groups. Multimodal integration can therefore improve prediction, but its value depends on sample size, missingness, modeling strategy, and the strength of the association between the available modalities and the clinical endpoint.

Model interpretation identified biologically coherent features that reproduced as prognostic biomarkers in PASS-01. *KDM6A* alterations were associated with inferior survival, consistent with human and experimental studies linking *KDM6A* loss to the squamous PDAC subtype(35,36), adverse prognosis(37), epigenetic reprogramming(37), tumor-cell plasticity(38), mTORC1 activation(39), and neutrophil recruitment(40). High *SSTR1* expression was associated with longer survival in both COMPASS and PASS-01. *SSTR1* is downregulated in pancreatic cancers, and experimental restoration suppresses proliferation, induces G0/G1 arrest, and limits xenograft growth(41). Co-expression with *SSTR2* also increases responsiveness to somatostatin analogs(42). The independent validation of *KDM6A* alteration and *SSTR1* expression establishes them as reproducible prognostic biomarkers and prioritizes these biologically distinct disease states for additional mechanistic study and therapeutic development. More broadly, the independent validation of model-derived features demonstrates that interpretable multimodal learning can extend beyond risk prediction to identify reproducible disease states and generate testable biological and therapeutic hypotheses.

This study has several limitations. First, although PASS-01 represents the largest randomized multimodal dataset available for metastatic PDAC, it remains a phase II trial and was not powered specifically for biomarker interaction testing. Consequently, confidence intervals for treatment-benefit estimates remain wide. Second, PFS was unavailable in COMPASS, requiring derivation of an exploratory PFS score rather than direct model training. Third, incomplete multimodal profiling necessitated late-fusion modeling to maximize patient inclusion. Fourth, PASS-01 excluded known or suspected germline *BRCA1*, *BRCA2*, and *PALB2* pathogenic variants, so these results do not extend to those subgroups. Finally, although external validation in PASS-01 substantially strengthens confidence in the findings, additional randomized cohorts will ultimately be required before biomarker-guided treatment selection can enter clinical practice.

In summary, multimodal pretreatment data contain reproducible prognostic information in metastatic PDAC, but predicting differential benefit between first-line regimens remains an open problem that will remain relevant as we enter an era with novel combinations on chemotherapy backbones. The PASS-01 Challenge provides a rigorous randomized benchmark to accelerate biomarker development for chemotherapy selection and represents an important step toward evidence-based precision treatment selection in pancreatic cancer.

## Data Availability

All data produced in the present study are available upon reasonable request to the authors

https://hpb-research.ca

## Acknowledgements

The authors used ChatGPT GPT-5.6 to generate and edit portions of this manuscript. All generated content was reviewed and approved by the authors.

This work was funded by the Princess Margaret Cancer Foundation, Ontario Institute for Cancer Research, University of Toronto Hold’em for Life, and the Marathon of Hope with the Terry Fox Research Institute.

## References

1. Rahib L, Smith BD, Aizenberg R, Rosenzweig AB, Fleshman JM, Matrisian LM. Projecting cancer incidence and deaths to 2030: the unexpected burden of thyroid, liver, and pancreas cancers in the United States. Cancer Res. 2014;74:2913–21.

2. Conroy T, Pfeiffer P, Vilgrain V, Lamarca A, Seufferlein T, O’Reilly EM, et al. Pancreatic cancer: ESMO Clinical Practice Guideline for diagnosis, treatment and follow-up. Ann Oncol. Elsevier BV; 2023;34:987–1002.

3. Guidelines Detail [Internet]. NCCN. [cited 2025 Nov 10]. Available from: https://www.nccn.org/guidelines/guidelines-detail?category=1&id=1455

4. Wainberg ZA, Melisi D, Macarulla T, Pazo Cid R, Chandana SR, De La Fouchardière C, et al. NALIRIFOX versus nab-paclitaxel and gemcitabine in treatment-naive patients with metastatic pancreatic ductal adenocarcinoma (NAPOLI 3): a randomised, open-label, phase 3 trial. Lancet. 2023;402:1272–81.

5. Knox JJ, O’Kane G, King D, Laheru D, Habowski AN, Yu K, et al. PASS-01: Randomized phase II trial of modified FOLFIRINOX versus gemcitabine/nab-paclitaxel and molecular correlatives for previously untreated metastatic pancreatic cancer. J Clin Oncol. 2025;JCO2500436.

6. Ohba A, Ozaka M, Mizusawa J, Okusaka T, Kobayashi S, Yamashita T, et al. Modified fluorouracil, leucovorin, irinotecan, and oxaliplatin or S-1, irinotecan, and oxaliplatin versus nab-paclitaxel + gemcitabine in metastatic or recurrent pancreatic cancer (GENERATE, JCOG1611): A randomized, open-label, phase II/III trial. J Clin Oncol. American Society of Clinical Oncology (ASCO); 2025;43:3345–54.

7. Nichetti F, Rota S, Ambrosini P, Pircher C, Gusmaroli E, Droz Dit Busset M, et al. NALIRIFOX, FOLFIRINOX, and gemcitabine with nab-paclitaxel as first-line chemotherapy for metastatic pancreatic cancer: A systematic review and meta-analysis: A systematic review and meta-analysis. JAMA Netw Open. American Medical Association (AMA); 2024;7:e2350756.

8. O’Kane GM, Grünwald BT, Jang G-H, Masoomian M, Picardo S, Grant RC, et al. GATA6 expression distinguishes classical and basal-like subtypes in advanced pancreatic cancer. Clin Cancer Res. American Association for Cancer Research (AACR); 2020;26:4901–10.

9. Wenric S, Sangli C, Guittar J, Islam F, Zander A, Won Hyun S, et al. Real-world validation of the purity independent subtyping of tumors classifier for informing therapy selection in pancreatic ductal adenocarcinoma. JCO Precis Oncol. American Society of Clinical Oncology (ASCO); 2025;9:e2500197.

10. Singh H, Xiu J, Kapner KS, Yuan C, Narayan RR, Oberley M, et al. Clinical and genomic features of classical and basal transcriptional subtypes in pancreatic cancer. Clin Cancer Res. American Association for Cancer Research (AACR); 2024;30:4932–42.

11. Rashid NU, Peng XL, Jin C, Moffitt RA, Volmar KE, Belt BA, et al. Purity Independent Subtyping of Tumors (PurIST), A clinically robust, single-sample classifier for tumor subtyping in pancreatic cancer. Clin Cancer Res. American Association for Cancer Research (AACR); 2020;26:82–92.

12. Perera S, Jang GH, Wang Y, Kelly D, Allen M, Zhang A, et al. HENT1 expression predicts response to gemcitabine and nab-paclitaxel in advanced pancreatic ductal adenocarcinoma. Clin Cancer Res. American Association for Cancer Research (AACR); 2022;28:5115–20.

13. Knox JJ, Jang GH, Grant RC, Zhang A, Ma L, Elimova E, et al. Whole genome and transcriptome profiling in advanced pancreatic cancer patients on the COMPASS trial. Nat Commun. Springer Science and Business Media LLC; 2025;16:5919.

14. Collins GS, Moons KGM, Dhiman P, Riley RD, Beam AL, Van Calster B, et al. TRIPOD+AI statement: updated guidance for reporting clinical prediction models that use regression or machine learning methods. BMJ. BMJ; 2024;385:e078378.

15. Aung KL, Fischer SE, Denroche RE, Jang G-H, Dodd A, Creighton S, et al. Genomics-driven precision medicine for advanced pancreatic cancer: Early results from the COMPASS trial. Clin Cancer Res. American Association for Cancer Research (AACR); 2018;24:1344–54.

16. Ter Veer E, van Rijssen LB, Besselink MG, Mali RMA, Berlin JD, Boeck S, et al. Consensus statement on mandatory measurements in pancreatic cancer trials (COMM-PACT) for systemic treatment of unresectable disease. Lancet Oncol. Elsevier BV; 2018;19:e151–60.

17. Maurer C, Holmstrom SR, He J, Laise P, Su T, Ahmed A, et al. Experimental microdissection enables functional harmonisation of pancreatic cancer subtypes. Gut. BMJ; 2019;68:1034–43.

18. Chan-Seng-Yue M, Kim JC, Wilson GW, Ng K, Figueroa EF, O’Kane GM, et al. Transcription phenotypes of pancreatic cancer are driven by genomic events during tumor evolution. Nat Genet. Springer Science and Business Media LLC; 2020;52:231–40.

19. Hanna MG, Reuter VE, Hameed MR, Tan LK, Chiang S, Sigel C, et al. Whole slide imaging equivalency and efficiency study: experience at a large academic center. Mod Pathol. Elsevier BV; 2019;32:916–28.

20. Xu H, Usuyama N, Bagga J, Zhang S, Rao R, Naumann T, et al. A whole-slide foundation model for digital pathology from real-world data. Nature. Springer Science and Business Media LLC; 2024;630:181–8.

21. Zou H, Hastie T. Regularization and variable selection via the elastic net. J R Stat Soc Series B Stat Methodol. Oxford University Press (OUP); 2005;67:301–20.

22. Chen T, Guestrin C. XGBoost: A Scalable Tree Boosting System [Internet]. arXiv [cs.LG]. 2016. Available from: 10.48550/arXiv.1603.02754

23. Hollmann N, Müller S, Eggensperger K, Hutter F. TabPFN: A Transformer that solves small tabular classification problems in a second [Internet]. arXiv [cs.LG]. 2022. Available from: 10.48550/arXiv.2207.01848

24. Hollmann N, Müller S, Purucker L, Krishnakumar A, Körfer M, Hoo SB, et al. Accurate predictions on small data with a tabular foundation model. Nature. Springer Science and Business Media LLC; 2025;637:319–26.

25. van Klaveren D, Steyerberg EW, Serruys PW, Kent DM. The proposed “concordance-statistic for benefit” provided a useful metric when modeling heterogeneous treatment effects. J Clin Epidemiol. Elsevier BV; 2018;94:59–68.

26. Golan T, O’Kane GM, Denroche RE, Raitses-Gurevich M, Grant RC, Holter S, et al. Genomic features and classification of homologous recombination deficient pancreatic ductal adenocarcinoma. Gastroenterology. Elsevier BV; 2021;160:2119–32.e9.

27. Davies H, Glodzik D, Morganella S, Yates LR, Staaf J, Zou X, et al. HRDetect is a predictor of BRCA1 and BRCA2 deficiency based on mutational signatures. Nat Med. Springer Science and Business Media LLC; 2017;23:517–25.

28. Roder J, Maguire L, Georgantas R 3rd, Roder H. Explaining multivariate molecular diagnostic tests via Shapley values. BMC Med Inform Decis Mak. 2021;21:211.

29. Wong A, Bourega TA, Nicolle R, Elqaderi A, Nowak KM, Light N, et al. Deep learning of histopathology predicts outcomes after surgery for pancreatic cancer. JCO Clin Cancer Inform. 2026;10:e2500268.

30. van Klaveren D, Maas CCHM, Kent DM. Measuring the performance of prediction models to personalize treatment choice: Defining observed and predicted pairwise treatment effects. Stat Med. 2023;42:4514–5.

31. Harrell FE Jr, Lee KL, Mark DB. Multivariable prognostic models: issues in developing models, evaluating assumptions and adequacy, and measuring and reducing errors. Stat Med. Wiley; 1996;15:361–87.

32. Alexandrov LB, Kim J, Haradhvala NJ, Huang MN, Tian Ng AW, Wu Y, et al. The repertoire of mutational signatures in human cancer. Nature. Springer Science and Business Media LLC; 2020;578:94–101.

33. Hendifar AE, Krishna V, Krishna V, Zhang H, Tarsode A, Nimgaonkar V, et al. Development and validation of a Computational Histology Artificial Intelligence-powered predictive biomarker for selection of chemotherapy in advanced pancreatic cancer. J Clin Oncol. American Society of Clinical Oncology (ASCO); 2026;JCO2502199.

34. O’Reilly EM, Lee JW, Zalupski M, Capanu M, Park J, Golan T, et al. Randomized, multicenter, phase II trial of gemcitabine and cisplatin with or without veliparib in patients with pancreas adenocarcinoma and a germline BRCA/PALB2 mutation. J Clin Oncol. American Society of Clinical Oncology (ASCO); 2020;38:1378–88.

35. Bailey P, Chang DK, Nones K, Johns AL, Patch A-M, Gingras M-C, et al. Genomic analyses identify molecular subtypes of pancreatic cancer. Nature. Springer Science and Business Media LLC; 2016;531:47–52.

36. Andricovich J, Perkail S, Kai Y, Casasanta N, Peng W, Tzatsos A. Loss of KDM6A activates super-enhancers to induce gender-specific squamous-like pancreatic cancer and confers sensitivity to BET inhibitors. Cancer Cell. Elsevier BV; 2018;33:512–26.e8.

37. Watanabe S, Shimada S, Akiyama Y, Ishikawa Y, Ogura T, Ogawa K, et al. Loss of KDM6A characterizes a poor prognostic subtype of human pancreatic cancer and potentiates HDAC inhibitor lethality. Int J Cancer. Wiley; 2019;145:192–205.

38. Yi Z, Wei S, Jin L, Jeyarajan S, Yang J, Gu Y, et al. KDM6A regulates cell plasticity and pancreatic cancer progression by noncanonical activin pathway. Cell Mol Gastroenterol Hepatol. Elsevier BV; 2022;13:643–67.

39. Revia S, Seretny A, Wendler L, Banito A, Eckert C, Breuer K, et al. Histone H3K27 demethylase KDM6A is an epigenetic gatekeeper of mTORC1 signalling in cancer. Gut. BMJ; 2022;71:1613–28.

40. Yang J, Jin L, Kim HS, Tian F, Yi Z, Bedi K, et al. KDM6A loss recruits tumor-associated neutrophils and promotes neutrophil extracellular trap formation in pancreatic cancer. Cancer Res. American Association for Cancer Research (AACR); 2022;82:4247–60.

41. Li M, Wang X, Li W, Li F, Yang H, Wang H, et al. Somatostatin receptor-1 induces cell cycle arrest and inhibits tumor growth in pancreatic cancer. Cancer Sci. Wiley; 2008;99:2218–23.

42. Li M, Zhang R, Li F, Wang H, Kim HJ, Becnel L, et al. Transfection of SSTR-1 and SSTR-2 inhibits Panc-1 cell proliferation and renders Panc-1 cells responsive to somatostatin analogue. J Am Coll Surg. Ovid Technologies (Wolters Kluwer Health); 2005;201:571–8.

