## Supplementary Tables and Figures for "Multimodal Machine Learning for Predicting Outcomes in the PASS-01 Trial of Systemic Therapy for Metastatic Pancreatic Cancer"

**Table S1: Cohort characteristics of patients in the COMPASS and PASS-01 trials, by treatment arm.**

| **Characteristic** | **COMPASS** | | **PASS-01** | |
| --- | --- | --- | --- | --- |
|  | **Modified**  **FOLFIRINOX** | **Gemcitabine/**  **nab-paclitaxel** | **Modified**  **FOLFIRINOX** | **Gemcitabine/**  **nab-paclitaxel** |
| Total | 144 | 99 | 80 | 80 |
| Data availability |  |  |  |  |
| Clinical | 144 (100%) | 99 (100%) | 80 (100%) | 80 (100%) |
| Pathology images | 109 (76%) | 72 (73%) | 71 (89%) | 71 (89%) |
| Whole genome | 144 (100%) | 99 (100%) | 65 (81%) | 62 (78%) |
| RNAseq | 137 (95%) | 92 (93%) | 58 (73%) | 57 (71%) |
| Age (years), median (IQR) | 62 (56–67) | 67 (61.5–72) | 62 (56–71) | 66 (60–70) |
| Sex (Male) | 94 (65%) | 54 (55%) | 49 (61%) | 52 (65%) |
| Race |  |  |  |  |
| White | 110 (76%) | 76 (77%) | 46 (58%) | 56 (70%) |
| Asian | 23 (16%) | 18 (18%) | 15 (19%) | 7 (9%) |
| Black/African American | 6 (4%) | 2 (2%) | 5 (6%) | 10 (13%) |
| Unknown | 5 (3%) | 3 (3%) | 14 (18%) | 7 (9%) |
| ECOG |  |  |  |  |
| 0 | 55 (38%) | 21 (21%) | 33 (41%) | 47 (59%) |
| 1 | 89 (62%) | 67 (68%) | 47 (59%) | 33 (41%) |
| 2 | 0 (0%) | 1 (1%) | 0 (0%) | 0 (0%) |
| CA 19-9 at baseline, median (IQR) | 1869 (273.85–9555) | 986 (95–4747) | 2063 (260.125–7510.25) | 1957.5 (101.25–11442.5) |
| Objective response | 41 (28%) | 30 (30%) | 20 (25%) | 21 (26%) |
| Progression event | NA | NA | 68 (85%) | 68 (85%) |
| Death event | 135 (94%) | 97 (98%) | 63 (79%) | 53 (66%) |

*Table S1 reflects only patients treated with FFX or GNP, the fixed cohort used for MULTIPL model development. In COMPASS, two FFX-treated patients lacking RECIST-based response assessment were excluded from this cohort for consistency across endpoints. In PASS-01, Table S1 reports the intention-to-treat (ITT) population (N=160; 80 FFX, 80 GNP). Fig. 1A reports data availability among all patients with the given modality profiled in COMPASS/PASS-01, regardless of treatment received, and therefore includes a broader population than the FFX/GNP modeling cohort. Fig. 1B is restricted to FFX/GNP-treated patients but reports evaluability separately for each endpoint; the two COMPASS patients excluded from Table S1 had valid one-year OS follow-up and are therefore included in the one-year OS evaluable count in Fig. 1B despite being excluded from Table S1 and from model development.*

**Table S2: Univariate survival analysis of top DNA features identified using SHAP for the model predicting one-year survival in the modified FOLFIRINOX arm of COMPASS.**

|  | **COMPASS Discovery** | | | **PASS-01 Validation** | | |
| --- | --- | --- | --- | --- | --- | --- |
| **Feature** | **N (high vs low)** | **HR (95% CI)** | **FDR** | **N (high vs low)** | **HR (95% CI)** | **P-value** |
| DSBR score | 131 vs 13 | 0.63 (0.35–1.12) | 0.315 | — | — | — |
| Deletion count | 72 vs 72 | 0.85 (0.61–1.20) | 0.653 | — | — | — |
| Insertion count | 72 vs 72 | 0.92 (0.66–1.30) | 0.867 | — | — | — |
| Signature 3 | 25 vs 119 | 0.50 (0.31–0.79) | 0.031 | 20 vs 107 | 0.77 (0.46–1.28) | 0.314 |
| Signature 13 | 72 vs 72 | 1.10 (0.78–1.54) | 0.867 | — | — | — |
| Signature 18 | 43 vs 101 | 1.53 (1.06–2.21) | 0.078 | 40 vs 87 | 0.79 (0.52–1.20) | 0.266 |
| Signature 2 | 72 vs 72 | 1.03 (0.73–1.45) | 0.867 | — | — | — |
| Signature 1 | 72 vs 72 | 1.04 (0.74–1.46) | 0.867 | — | — | — |
| Signature 5 | 72 vs 72 | 1.06 (0.76–1.49) | 0.867 | — | — | — |
| *KDM6A* alterations | 11 vs 133 | 2.21 (1.19–4.11) | 0.056 | 12 vs 115 | 2.71 (1.46–5.00) | <0.001 |
| Signature 17 | 72 vs 72 | 1.23 (0.88–1.72) | 0.508 | — | — | — |
| *Continuous DNA features were dichotomized at the COMPASS median into high and low groups; KDM6A alteration status was used directly as a binary feature (mutated vs. wild-type). Features meeting the false-discovery rate (FDR) < 0.1 threshold in COMPASS discovery were carried forward for PASS-01 validation; PASS-01 results are not shown for features not meeting this threshold. HR = hazard ratio; CI = confidence interval; FDR = false-discovery rate; DSBR = Double-Strand Break Repair. PASS-01 p-values are unadjusted.* | | | | | | |

**Table S3: Univariate survival analysis of top RNA features identified using SHAP for the model predicting one-year survival in the modified FOLFIRINOX arm of COMPASS.**

|  | **COMPASS Discovery** | | | **PASS-01 Validation** | | |
| --- | --- | --- | --- | --- | --- | --- |
| **Gene** | **N (high vs low)** | **HR (95% CI)** | **FDR** | **N (high vs low)** | **HR (95% CI)** | **P-value** |
| *DDR2* | 69 vs 68 | 0.57 (0.40–0.80) | 0.010 | 97 vs 18 | 0.74 (0.43–1.27) | 0.276 |
| *ELAC2* | 69 vs 68 | 1.38 (0.97–1.95) | 0.082 | — | — | — |
| *SNCB* | 32 vs 105 | 0.67 (0.44–1.00) | 0.070 | 24 vs 91 | 1.20 (0.74–1.93) | 0.465 |
| *MYO15A* | 69 vs 68 | 0.68 (0.48–0.97) | 0.068 | 53 vs 62 | 0.80 (0.53–1.20) | 0.276 |
| *RBM10* | 69 vs 68 | 1.25 (0.88–1.77) | 0.210 | — | — | — |
| *TPTEP2-CSNK1E* | 69 vs 68 | 0.70 (0.50–1.00) | 0.070 | 37 vs 78 | 0.93 (0.60–1.42) | 0.727 |
| *FAM131B* | 69 vs 68 | 0.69 (0.49–0.98) | 0.068 | 84 vs 31 | 0.94 (0.59–1.49) | 0.778 |
| *WRAP53* | 69 vs 68 | 1.62 (1.14–2.30) | 0.025 | 107 vs 8 | 0.70 (0.30–1.62) | 0.401 |
| *UFM1* | 69 vs 68 | 0.73 (0.51–1.03) | 0.082 | — | — | — |
| *AC087289.3* | 69 vs 68 | 1.45 (1.03–2.06) | 0.068 | 71 vs 44 | 0.96 (0.64–1.45) | 0.850 |
| *PRSS3* | 69 vs 68 | 0.58 (0.41–0.82) | 0.010 | 80 vs 35 | 0.82 (0.54–1.26) | 0.365 |
| *All expression levels were dichotomized at the COMPASS median into high and low groups, except SNCB, which was dichotomized as present vs. absent (median = 0). Features meeting the false-discovery rate (FDR) < 0.1 threshold in COMPASS discovery were carried forward for PASS-01 validation; PASS-01 results are not shown for features not meeting this threshold. ELAC2 and UFM1 met this threshold but were not validated in PASS-01 because no samples were in the low-expression category after applying the COMPASS-derived threshold. HR = hazard ratio; CI = confidence interval; FDR = false-discovery rate. PASS-01 p-values are unadjusted.* | | | | | | |

**Table S4: Univariate survival analysis of top DNA features identified using SHAP for the model predicting one-year survival in the gemcitabine/nab-paclitaxel arm of COMPASS.**

|  | **COMPASS Discovery** | | | **PASS-01 Validation** | | |
| --- | --- | --- | --- | --- | --- | --- |
| **Feature** | **N (high vs low)** | **HR (95% CI)** | **FDR** | **N (high vs low)** | **HR (95% CI)** | **P-value** |
| MMR score | 56 vs 43 | 1.66 (1.10–2.51) | 0.057 | 74 vs 53 | 1.38 (0.94–2.03) | 0.100 |
| *SPEN* alterations | 4 vs 95 | 0.40 (0.15–1.11) | 0.140 | — | — | — |
| Deletion count | 50 vs 49 | 1.13 (0.76–1.68) | 0.559 | — | — | — |
| SNV count | 50 vs 49 | 0.75 (0.50–1.12) | 0.212 | — | — | — |
| *Continuous DNA features were dichotomized at the COMPASS median into high and low groups; SPEN alteration status was used directly as a binary feature. Features meeting the false-discovery rate (FDR) < 0.1 threshold in COMPASS discovery were carried forward for PASS-01 validation; PASS-01 results are not shown for features not meeting this threshold. HR = hazard ratio; CI = confidence interval; FDR = false-discovery rate; MMR = mismatch repair; SNV = single nucleotide variant. PASS-01 p-values are unadjusted.* | | | | | | |

**Table S5: Univariate survival analysis of top RNA features identified using SHAP for the model predicting one-year survival in the gemcitabine/nab-paclitaxel arm of COMPASS.**

|  | **COMPASS Discovery** | | | **PASS-01 Validation** | | |
| --- | --- | --- | --- | --- | --- | --- |
| **Gene** | **N (high vs low)** | **HR (95% CI)** | **FDR** | **N (high vs low)** | **HR (95% CI)** | **P-value** |
| *SEMA3A* | 46 vs 46 | 2.20 (1.41–3.42) | 0.001 | 81 vs 34 | 1.17 (0.75–1.81) | 0.490 |
| *HUNK* | 46 vs 46 | 0.92 (0.60–1.39) | 0.678 | — | — | — |
| *AL117339.4* | 46 vs 46 | 0.45 (0.29–0.70) | 0.001 | 98 vs 17 | 0.77 (0.44–1.36) | 0.367 |
| *AC098484.3* | 46 vs 46 | 0.56 (0.36–0.86) | 0.014 | 53 vs 62 | 1.24 (0.83–1.84) | 0.300 |
| *MTCP1* | 46 vs 46 | 0.39 (0.25–0.62) | <0.001 | 109 vs 6 | 0.88 (0.32–2.40) | 0.797 |
| *FAM53C* | 46 vs 46 | 0.78 (0.51–1.18) | 0.264 | — | — | — |
| *PDE3B* | 46 vs 46 | 0.47 (0.30–0.72) | 0.001 | 83 vs 32 | 0.80 (0.51–1.24) | 0.308 |
| *SEMA6A* | 46 vs 46 | 0.71 (0.47–1.09) | 0.146 | — | — | — |
| *KDM4A* | 46 vs 46 | 0.48 (0.31–0.74) | 0.002 | — | — | — |
| *SSTR1* | 46 vs 46 | 0.59 (0.39–0.90) | 0.020 | 68 vs 47 | 0.59 (0.39–0.88) | 0.009 |
| *All expression levels were dichotomized at the COMPASS median into high and low groups. Features meeting the false-discovery rate (FDR) < 0.1 threshold in COMPASS discovery were carried forward for PASS-01 validation; PASS-01 results are not shown for features not meeting this threshold. KDM4A met this threshold but was not validated in PASS-01 because no samples were in the low-expression category after applying the COMPASS-derived threshold. HR = hazard ratio; CI = confidence interval; FDR = false-discovery rate. PASS-01 p-values are unadjusted.* | | | | | | |

**Table S6: Data provided through the PASS-01 Challenge and variables included in modeling.**

| **Modality** | **Variable/Data** | **Provided in PASS-01 Challenge Data** | **Included in MULTIPL Model** |
| --- | --- | --- | --- |
| Clinical | Age | Yes | Yes |
|  | Gender | Yes | Yes |
|  | Race | Yes | Yes |
|  | Baseline ECOG | Yes | Yes |
|  | Baseline CA 19-9 | Yes | Yes |
|  | Treatment arm | Yes | Yes - prognostic models only |
|  | Site of biopsy | Yes | No |
|  | Metastatic sites | Yes | No |
| DNA | Tumor and matched normal genomic CRAM files | Yes | Yes - Panel gene binary mutation status; mutational signatures |
| RNA | Tumor transcriptomic BAM files | Yes | Yes - Protein-coding gene expression (RNA-seq) |
| Histopathology | Digitized histopathology slide images | Yes | Yes - GigaPath embeddings |

### Supplementary Figures

**Figure S1. Development of MULTIPL for objective response prediction in COMPASS.** **A–B**, Cross-validated performance of candidate models in patients treated with FFX (**A**) and GNP (**B**). Points indicate area under the receiver operating characteristic curve; horizontal lines indicate 95% bootstrap confidence intervals. Models are grouped by fusion strategy and unimodal data type. For late-fusion models, combinations of base-learner and meta-learner are shown. **C–D**, Best percentage change in target-lesion size in patients treated with FFX (**C**) and GNP (**D**). The bar color indicates the predicted probability of an objective response from the selected late-fusion model. Dashed lines indicate RECIST 1.1 thresholds for progressive disease and partial response; non-evaluable patients are shown separately. AUC, area under the receiver operating characteristic curve; CLIN, clinical; HIST, histopathology; FFX, modified FOLFIRINOX; GNP, gemcitabine plus nab-paclitaxel.

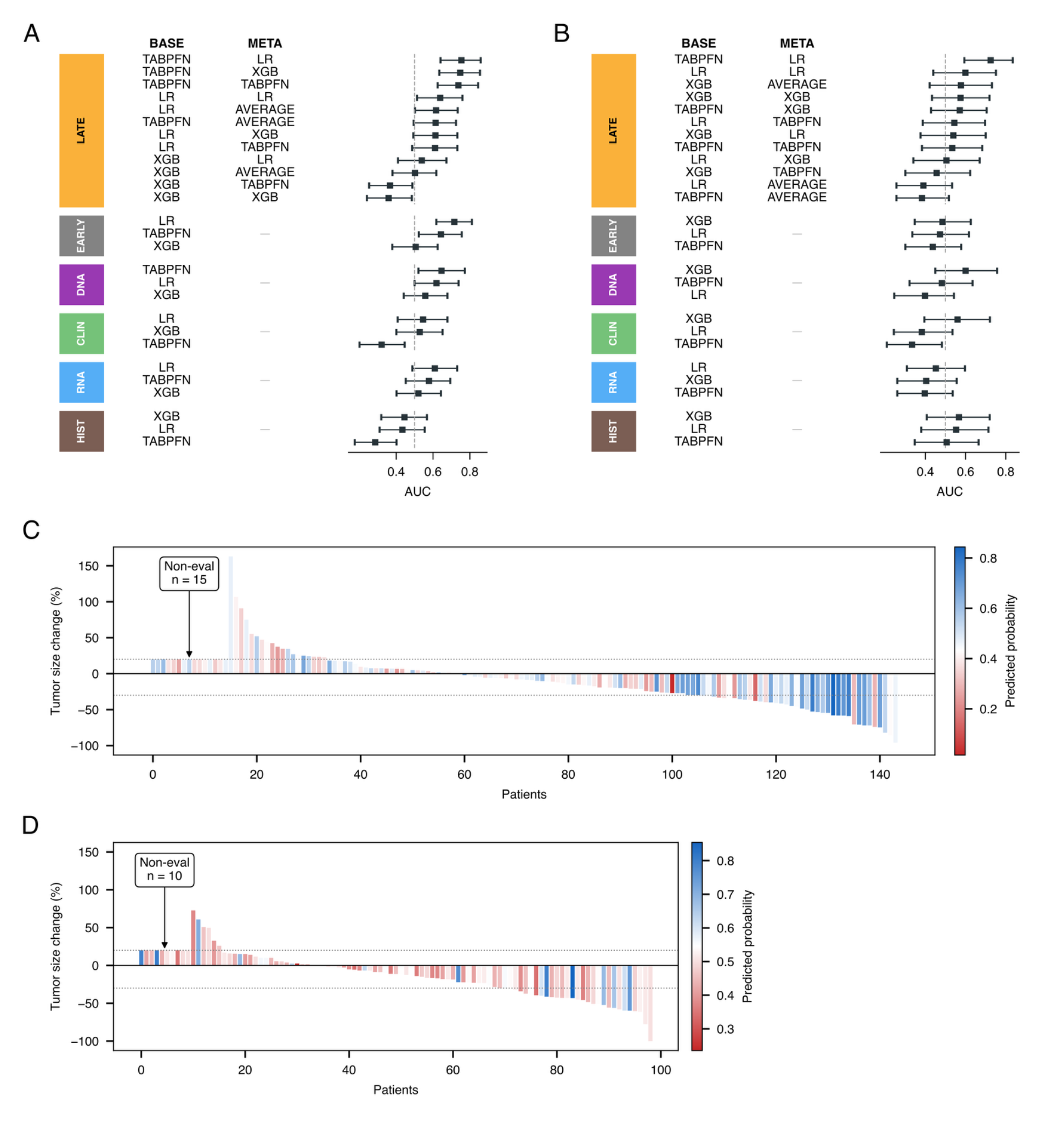

**Figure S2. Correlation of unimodal and multimodal predictions in COMPASS.** **A–B**, Pairwise Pearson correlations among predicted one-year overall survival probabilities from unimodal clinical, DNA, RNA, and histopathology TabPFN models, an early-fusion TabPFN model, and the selected late-fusion model in the FFX (**A**) and GNP (**B**) arms. **C–D**, Corresponding correlations for objective-response predictions in the FFX (**C**) and GNP (**D**) arms. Cell values indicate Pearson correlation coefficients; colors denote the direction and magnitude. FFX, modified FOLFIRINOX; GNP, gemcitabine plus nab-paclitaxel.

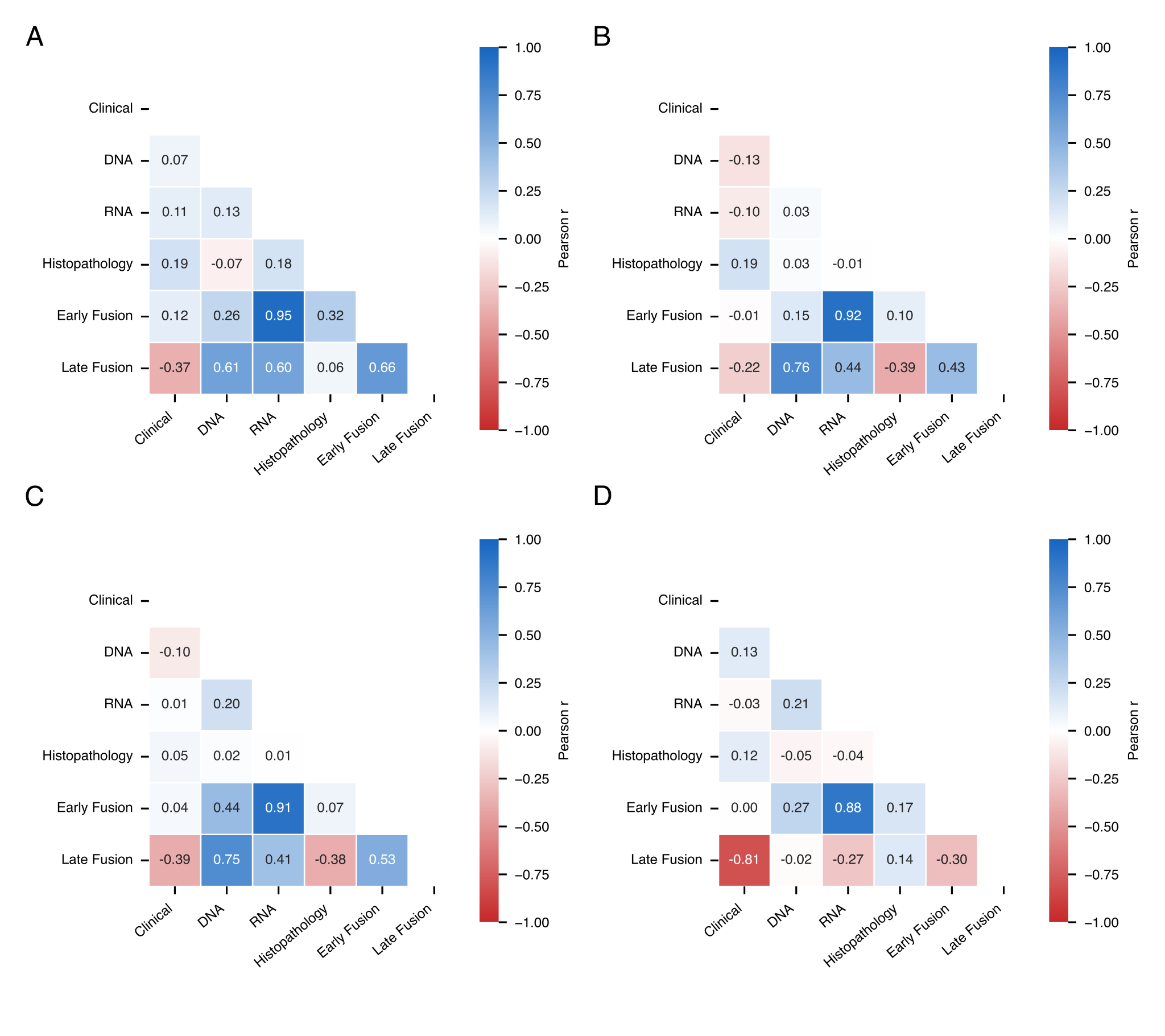

**Figure S3. Evaluation of MULTIPL in COMPASS using the PASS-01 Challenge framework.** **A, C**, Differential treatment effects analyses for overall survival (**A**) and objective response (**C**). Treatment-effect scores were calculated as the predicted outcome with FFX minus the predicted outcome with GNP; positive values favor FFX. Kaplan–Meier curves are stratified by observed treatment within model-defined treatment-recommendation groups. **B, D**, Prognostic analyses for overall survival (**B**) and objective response (**D**). Risk groups were defined using the median. Hazard ratios compare GNP with FFX in treatment-effect analyses and high-risk with low-risk in prognostic analyses. PFS was unavailable in COMPASS. FFX, modified FOLFIRINOX; GNP, gemcitabine plus nab-paclitaxel; ORR, objective response; OS, overall survival.

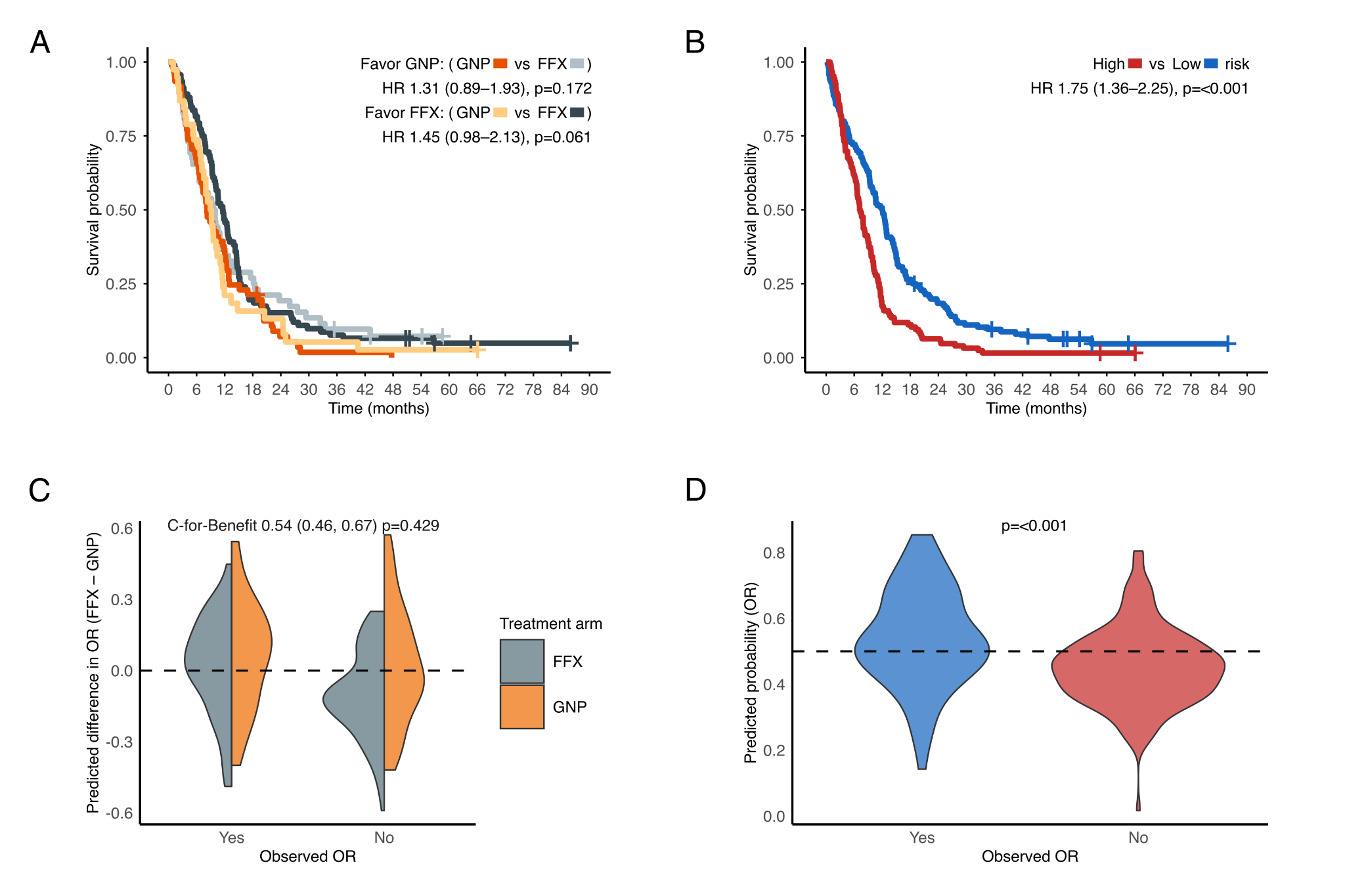

**Figure S4. PurIST performance in the PASS-01 Challenge.** **A, C, E**, Differential treatment effects analyses for overall survival (**A**), objective response (**C**), and progression-free survival (**E**). PurIST scores were derived from RNA expression and converted to treatment-effect scores, with classical cancers classified as “favor FFX” and basal-like cancers as “favor GNP”. Kaplan–Meier curves were stratified by randomized treatment within model-defined treatment-recommendation groups. **B, D, F**, Prognostic analyses with basal-like cancers classified as high risk and classical cancers as low risk for overall survival (**B**), objective response (**D**), and progression-free survival (**F**). FFX, modified FOLFIRINOX; GNP, gemcitabine plus nab-paclitaxel; HR, hazard ratio; ORR, objective response.

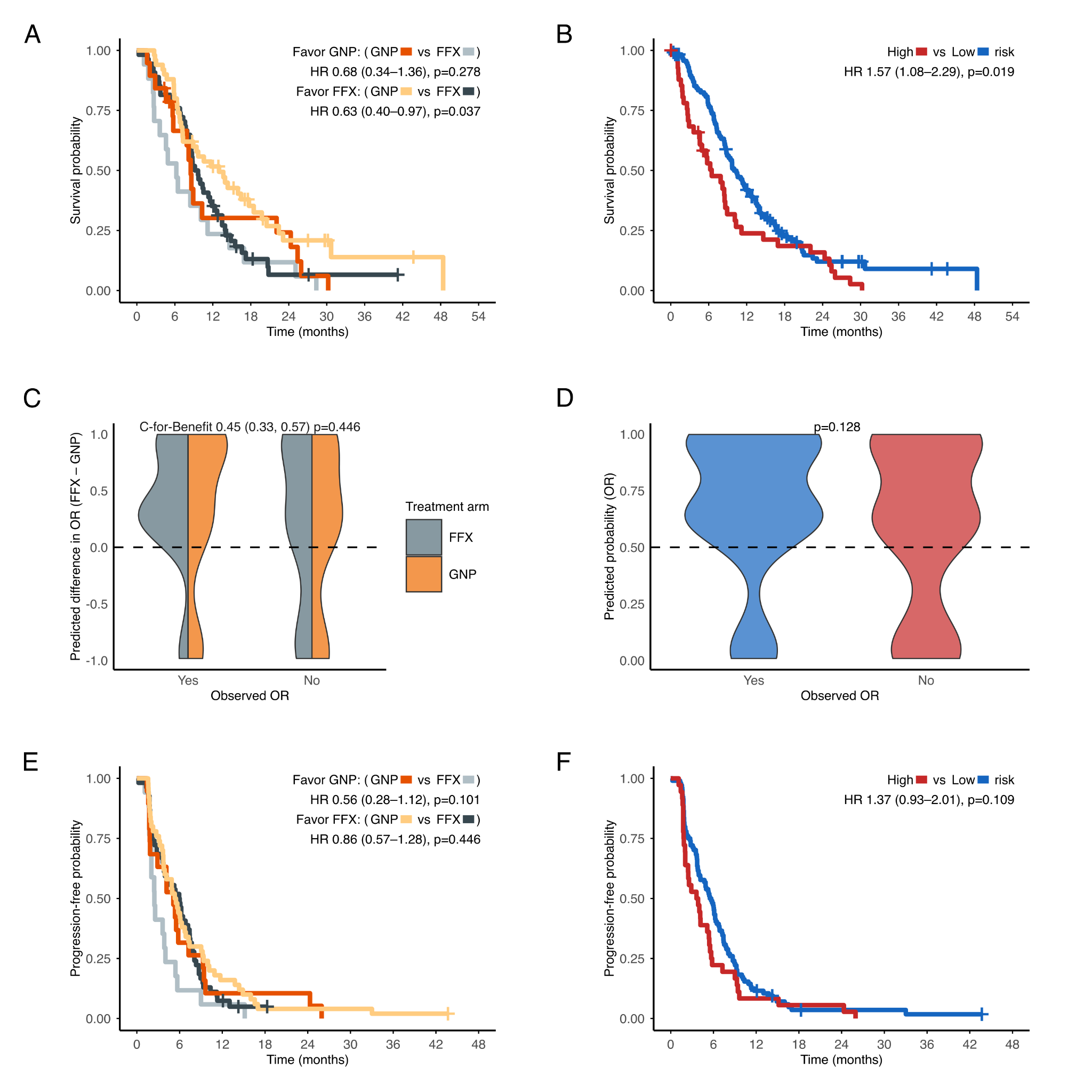

**Figure S5. hENT1 performance in the PASS-01 Challenge.** **A, C, E**, Differential treatment effects analyses for overall survival (**A**), objective response (**C**), and progression-free survival (**E**). High hENT1 expression, defined as the median in COMPASS, was classified as “favor GNP”. Kaplan–Meier curves were stratified by randomized treatment within model-defined treatment-recommendation groups. **B, D, F**, Prognostic analyses comparing hENT1 high versus low based on the COMPASS median for overall survival (**B**), objective response (**D**), and progression-free survival (**F**). The GNP-favoring and hENT1-high groups were small, limiting precision. FFX, modified FOLFIRINOX; GNP, gemcitabine plus nab-paclitaxel; HR, hazard ratio; ORR, objective response.

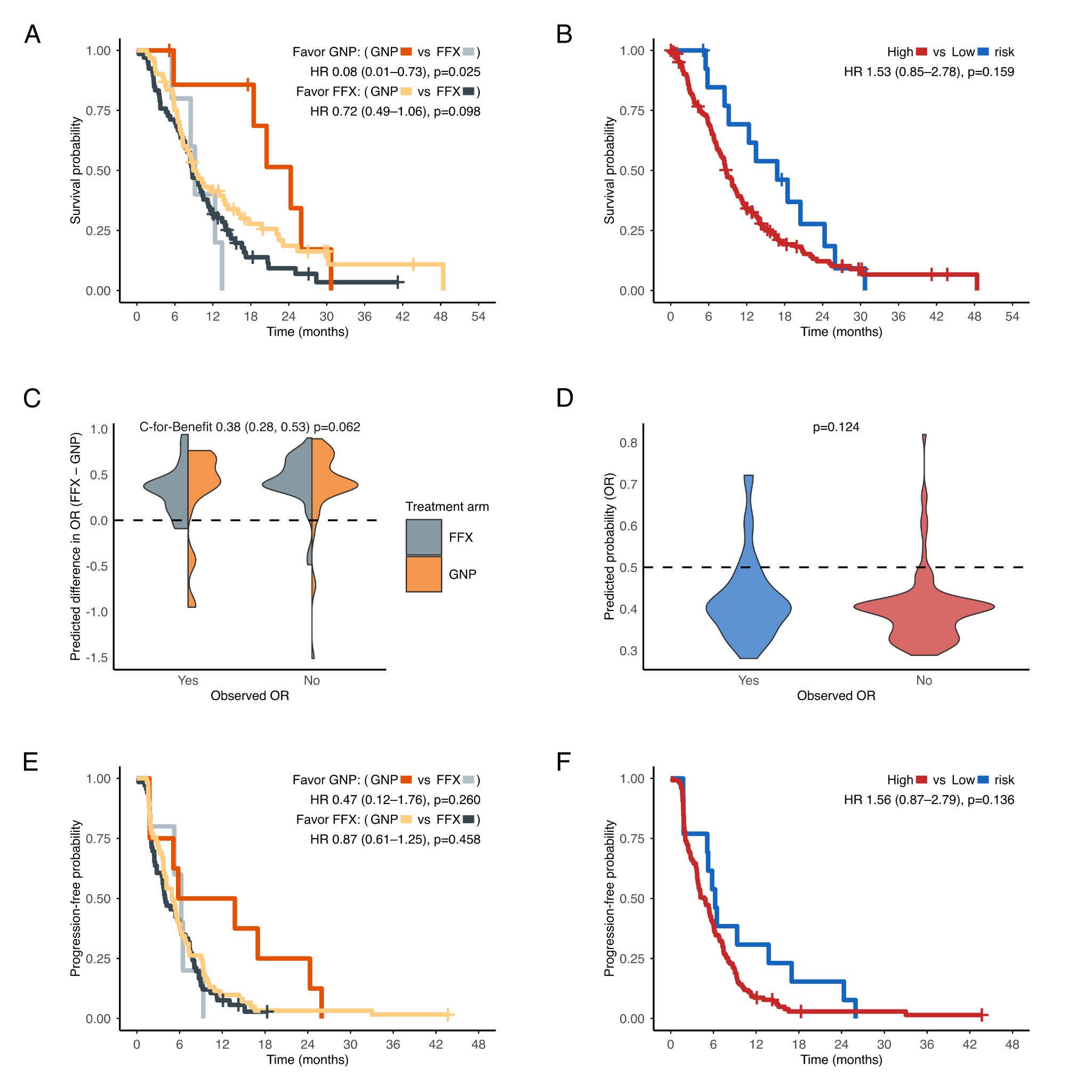

**Figure S6. HRDetect performance in the PASS-01 Challenge.** **A, C, E**, Differential treatment effects analyses for overall survival (**A**), objective response (**C**), and progression-free survival (**E**). HRDetect scores were derived from whole-genome sequencing, and cancers above the published threshold of 0.7 were classified as “favor FFX”. Kaplan–Meier curves were stratified by randomized treatment within treatment-recommendation groups. **B, D, F**, Prognostic analyses with HRDetect over 0.7 as high risk for overall survival (**B**), objective response (**D**), and progression-free survival (**F**). Hazard ratios compare GNP with FFX in treatment-effect analyses and high-risk with low-risk in prognostic analyses. The FFX-favoring and HRD-high groups were small, limiting interpretation. FFX, modified FOLFIRINOX; GNP, gemcitabine plus nab-paclitaxel; HR, hazard ratio; ORR, objective response.

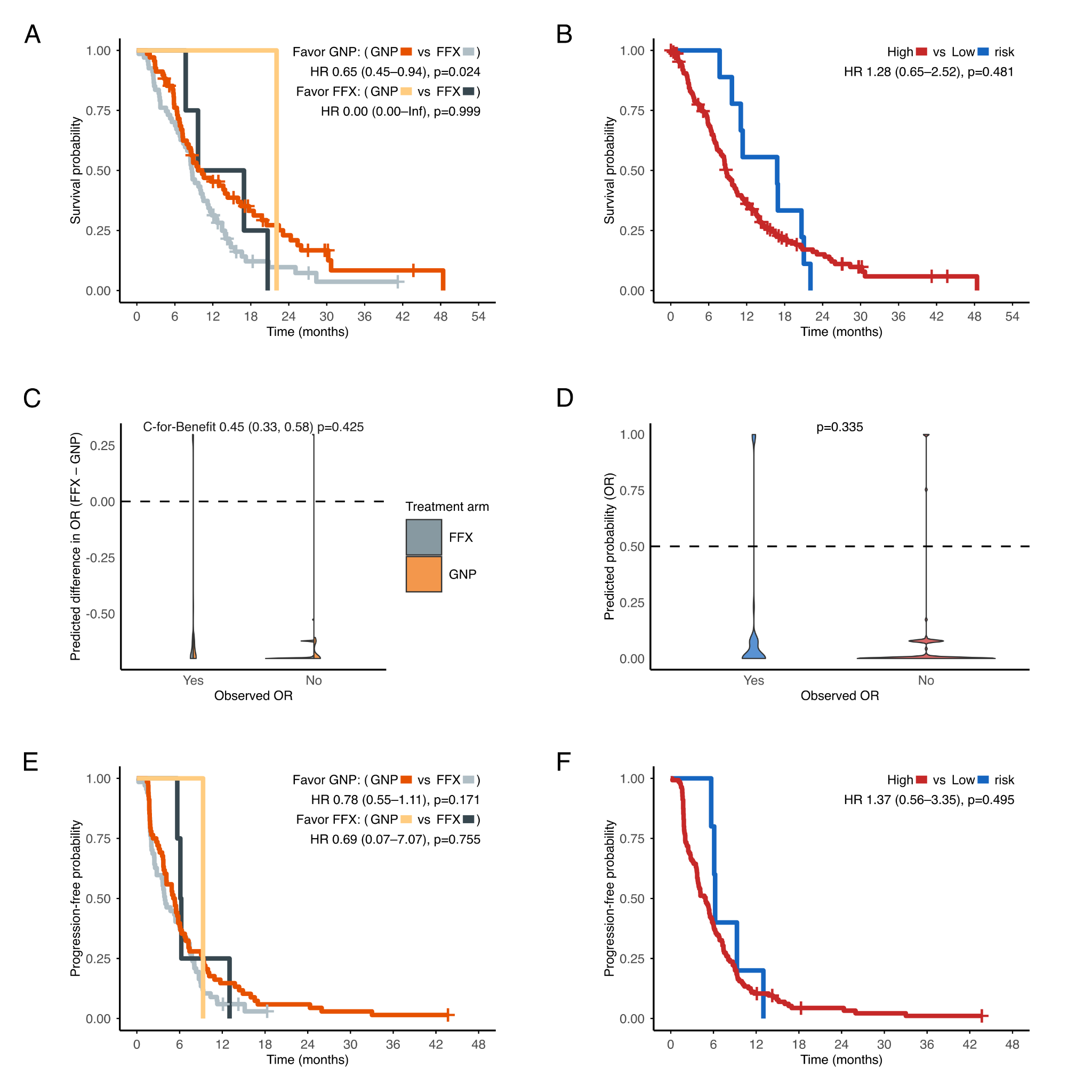

**Figure S7. Rank-average ensemble performance in the PASS-01 Challenge.** **A, C, E**, Differential treatment effects analyses for overall survival (**A**), objective response (**C**), and progression-free survival (**E**). Treatment-effect scores were defined as the rank-average predicted outcome with FFX minus that with GNP; positive values were classified as “favor FFX” and negative values as “favor GNP.” Kaplan–Meier curves were stratified by randomized treatment within model-defined treatment-recommendation groups. **B, D, F**, Prognostic analyses comparing ensemble-defined high- versus low-risk groups based on the COMPASS median for predicted overall survival (**B**), objective response (**D**), and progression-free survival (**F**). The PFS score was derived from one-year overall survival and objective-response model outputs. FFX, modified FOLFIRINOX; GNP, gemcitabine plus nab-paclitaxel; HR, hazard ratio; ORR, objective response.

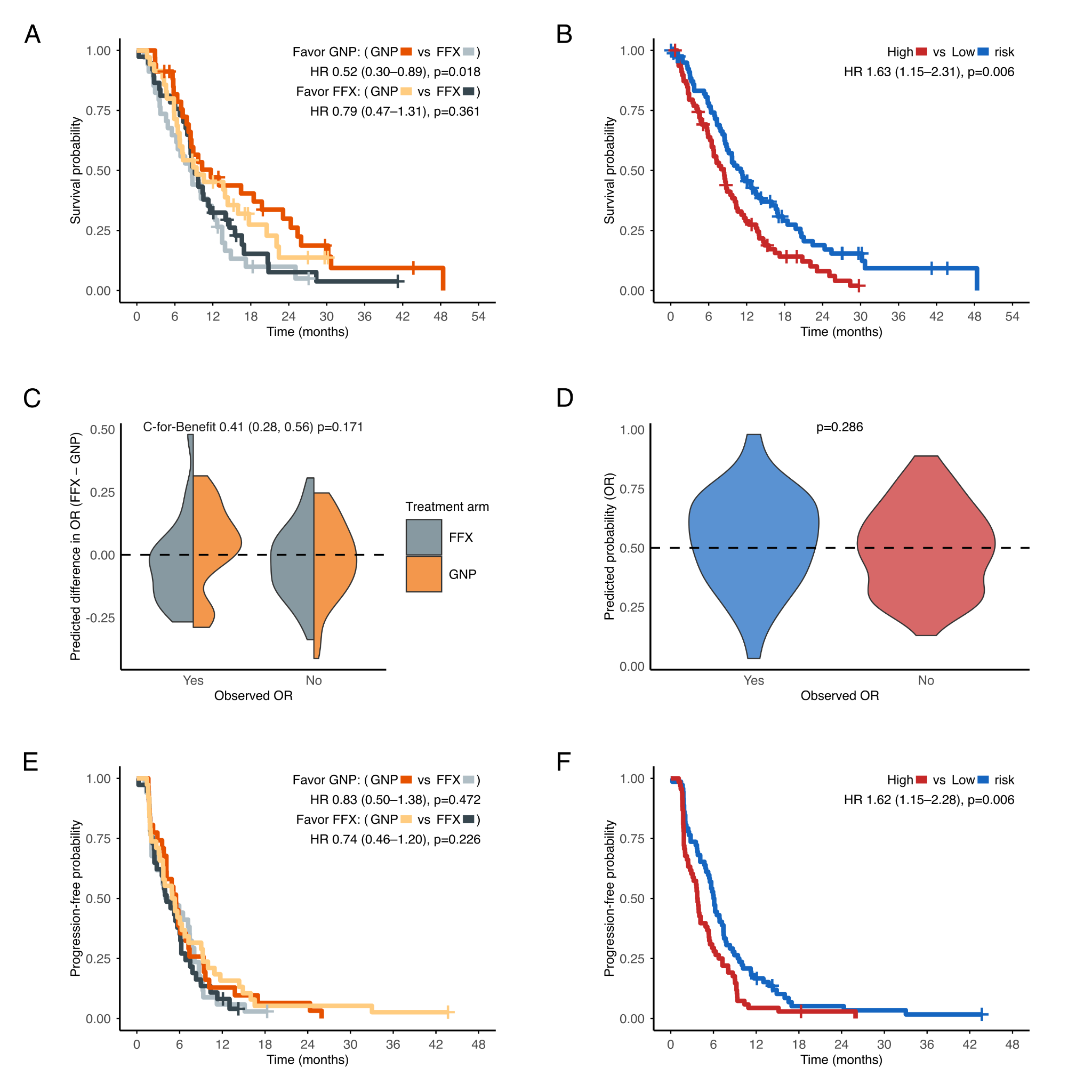
